# Insights into COVID-19 epidemiology and control from temporal changes in serial interval distributions in Hong Kong

**DOI:** 10.1101/2022.08.29.22279351

**Authors:** Sheikh Taslim Ali, Dongxuan Chen, Wey Wen Lim, Amy Yeung, Dillon C. Adam, Yiu Chung Lau, Eric H. Y. Lau, Jessica Y. Wong, Jingyi Xiao, Faith Ho, Huizhi Gao, Lin Wang, Xiao-Ke Xu, Zhanwei Du, Peng Wu, Gabriel M. Leung, Benjamin J. Cowling

**Affiliations:** WHO Collaborating Centre for Infectious Disease Epidemiology and Control, School of Public Health, Li Ka Shing Faculty of Medicine, The University of Hong Kong, Hong Kong Special Administrative Region, China; Laboratory of Data Discovery for Health Limited, Hong Kong Science and Technology Park, New Territories, Hong Kong; Department of Genetics, University of Cambridge, Cambridge, United Kingdom; College of Information and Communication Engineering, Dalian Minzu University, Dalian 116600, China

## Abstract

The serial interval distribution is used to approximate the generation time distribution, an essential parameter to predict the effective reproductive number “*R*_*t*_”, a measure of transmissibility. However, serial interval distributions may change as an epidemic progresses rather than remaining constant. Here we show that serial intervals in Hong Kong varied over time, closely associated with the temporal variation in COVID-19 case profiles and public health and social measures that were implemented in response to surges in community transmission. Quantification of the variation over time in serial intervals led to improved estimation of *R*_*t*_, and provided additional insights into the impact of public health measures on transmission of infections.

**One-Sentence Summary:** Real-time estimates of serial interval distributions can improve assessment of COVID-19 transmission dynamics and control.

## Main Text

Monitoring the intensity of coronavirus disease 2019 (COVID-19) transmission is an essential component of public health surveillance for situational awareness and real-time impact assessment of interventions (*1, 2*). Transmissibility has typically been measured by the effective reproductive number “*R*_*t*_” based on analysis of epidemic curves along with an estimate of the generation time distribution (*3-5*). The generation time for an infectious disease describes the average time between consecutive infections in a transmission chain (*4, 6*), and is often approximated by the serial interval distribution which describes the average time between illness onsets of consecutive cases in a transmission chain (*3, 7, 8*). However, we and others have shown that the serial interval distribution may change as an epidemic progresses rather than remaining constant (*9-11*).

In Hong Kong, a subtropical city located on the southern coast of China, the public health responses to COVID-19 following a “dynamic zero covid” strategy aiming to eliminate local infections included a number of key components. First, travel restrictions and on-arrival quarantines on all in-bound travellers have been implemented throughout the pandemic to reduce the importation risk. Second, a series of targeted and community-wide public health and social measures have been used to minimize local transmission of COVID-19, including hospital isolation of all confirmed cases, quarantine of close contacts in designated facilities, mask mandate, restriction on gatherings, closures of facilities, schools, and some workplaces, and compulsory testing orders for persons at a higher risk of exposure. This strategy was successful in limiting cumulative incidence to 12,600 confirmed cases by 31 December 2021, corresponding to 0.16% of the total population in Hong Kong. All community epidemics were associated with ancestral-like viruses and not variants. Here, our objective is to take advantage of detailed contact tracing data in Hong Kong to characterise temporal changes within and between waves in the serial interval distribution of COVID-19 through a series of community epidemics, and identify possible factors associated with those temporal changes.

### COVID-19 transmission and PHSMs in Hong Kong

We obtained detailed information on each laboratory-confirmed case of COVID-19 from the Department of Health of the Hong Kong SAR government (section 1, supplementary materials). The database included information on the confirmation date, illness onset date, isolation date, hospital admission date, outcome (critical/serious/stable), infection origin (locally infected vs infected outside Hong Kong) and arrival date (if applicable), home and workplace location, detailed travel/movement history, cluster information from contact tracing (if applicable) of each confirmed case (*12*). In Hong Kong from 22 January 2020 through 31 July 2022, COVID-19 has caused five local epidemic waves. The “first wave”, defined as the period from late January to mid-February 2020 with a small number of community cases linked to importations from mainland China (*13*). Given the very small number of imported infections (n=45) we excluded the first wave from our study. We analysed data from throughout the second wave (1 March 2020 – 10 April 2020), third wave (25 June 2020 – 8 September 2020) and fourth wave (1 Nov 2020 – 23 March 2021) (**Fig. 1A**). In the fifth wave which started in early January 2022 (*14*), contact tracing capacity was challenged due to the fast exponential rise in cases, and we were only able to include detailed data on confirmed transmission pairs from the very early stage of this wave in these analyses. Our study received ethical approval from the Institutional Review Board of the University of Hong Kong (ref: UW 20-341).

**Fig. 1.**
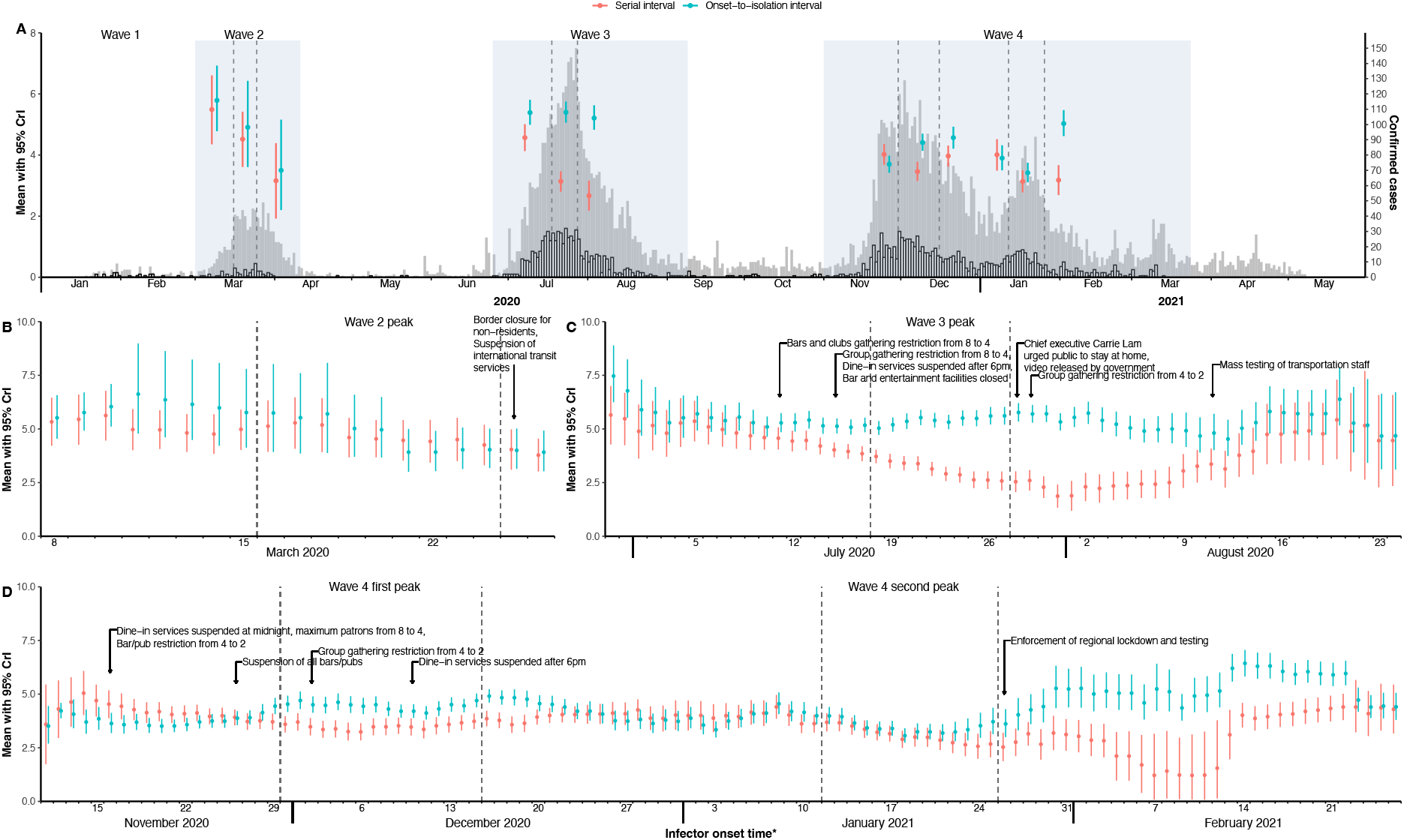
Transmission dynamics and temporal estimates of serial interval distribution and isolation delay for COVID-19 in Hong Kong. (A) The grey bars indicate the epi-curve of the reported COVID-19 cases, and white bars are onset epi-curves for infectors of those reported cases during four waves in Hong Kong. Mean serial interval estimates (red dots) with 95% CrI (red vertical line segments) and mean onset-to-isolation interval (teal dots) with 95% CrI (teal vertical line segments), evaluated from all confirmed and likely transmission pairs during pre-peak, peak-timing, and post-peaks for three pre-defined waves (light blue shades) with vertical dark grey dashed lines referring to the peak-timing for each wave, with two peaks for fourth wave. (B)-(D) Time varying estimates of effective serial intervals and onset-to-isolation interval for second wave (A), third wave (B) and fourth wave (C) with the indicator to timings of major public health and social measures (PHSMs) implemented in Hong Kong. The area between two grey dashed lines in each wave indicates the peak timing of the epidemic wave. The estimates of serial interval and onset-to-isolation interval are evaluated by using MCMC on fitting normal and gamma distributions to empirical data on confirmed and likely transmission pairs (considering predefined threshold of 8 days) respectively. The effective serial intervals and onset-to-isolation intervals are estimated based on the empirical data in 10-day sliding windows (presented with respect to the 5^th^ day of each sliding window).

We collected information from official reports on the public health and social measures (PHSMs) implemented in Hong Kong to control the spread of COVID-19 (*15*). We further classified these interventions into case-based, community-wide (including travel-based) control measures (*16*). The case-based measures including strict isolation of cases and quarantine of close contacts were maintained throughout each wave while the community-wide measures were generally only implemented when needed to bring each epidemic wave under control. The summary of these PHSMs including their timing and duration is provided in Supplementary table S1.

### Construction of transmission pairs

Following our earlier work (*17*), we first reconstructed the initial transmission pairs with reference to the detailed individual data on all confirmed cases retrieved from the Centre for Health Protection. We further rechecked the initially constructed pairs by their cluster information and epidemiological linkage with other cases and determined the infector and infectee within a pair according to a pre-defined algorithm (section 1, supplementary materials). The larger clusters with complex epidemiological linkages between the cases were individually assessed to determine the infector and the infectee in each transmission pair. If there were two or more likely infectors in one cluster, we defined the infector as the case with the earliest onset date. If cases shared the same onset date, the one with an earlier report date or case number would be classified as an infector (section 1, supplementary materials) (*9, 18*). We also performed a possible cross-check on unclear pairs with phylogenetic data to examine potential infector-infectee relationships with the *Phybreak* package in R (*19*). We resolved the multiple infector issue by identifying a case to be the infector of all subsequent cases with an immediate link in a cluster when the onset intervals between the infector and the infectees fell within a pre-defined time period, 8 days for the main analysis and 5-14 days for the sensitivity analyses (section 6, supplementary materials). Among more than 12,600 COVID-19 cases in Hong Kong in the period 1 March 2020 through to 23 March 2021, most were identified in the third and fourth waves (**Fig. 1A**), in total we were able to construct 2433 transmission pairs, including 87, 965 and 1381 in the second, third and fourth waves, respectively. Among these, we considered 47, 357 and 355 transmission pairs as “confirmed pairs” in these three waves, respectively, and the remainder were “likely pairs” (section 1, supplementary materials). There were an additional 64 pairs identified in small outbreaks occurred between our pre-defined waves, and 229 pairs (30 for BA1, 174 for BA2 and 25 for delta) identified in the very early stage of the fifth wave (fig. S1).

### Time-varying serial intervals and PHSMs

We estimated the serial interval distributions over time in a Bayesian framework based on Markov Chain Monte Carlo, implemented with the *RStan* package in R (section 2, supplementary materials). We estimated serial interval distributions for three periods within each wave i.e. pre-peak, during peak, and post-peak, and then estimated the time-varying daily effective serial interval distributions with a sliding window of 10 days (7-14 days for sensitivity analysis) (*9*). We used the same framework to estimate the distributions of onset-to-isolation intervals (indicating case isolation delay) for the infectors and then performed multivariable regression analysis on mean serial intervals where the mean onset-to-isolation interval and various PHSMs were included as explanatory variables (section 3, supplementary materials). The variation in population mobility and mixing, potentially reflecting the effect of these PHSMs, might also impact the transmission dynamics of COVID-19 (*20-22*). Therefore, we also performed a multivariable regression analysis on the estimated mean serial intervals with the daily digital transactions through Octopus cards, a widely-used contactless electronic payment system in Hong Kong, as the proxy of relative mobility of the population (*20*), and daily per capita testing volume (number of PCR tests conducted per 10,000 population per day) in Hong Kong (section 3, supplementary materials).

Among the 2497 transmission pairs identified in during the first two years of the COVID-19 pandemic in Hong Kong including the second, third and fourth waves with the ancestral strain of SARS-CoV-2, the mean serial interval (*μ*) was estimated to be 3.6 (95% CrI: 3.5, 3.7) days with standard deviation (*σ*) 3.4 (95% CrI: 3.3, 3.5) days.

We identified the peak of the second wave from 16 to 24 March 2020, from 18 to 27 July 2020 for the third wave, and the fourth wave had two peaks, the first from 30 November 2020 to 15 December 2020 and the second during 11 - 25 January 2021 (section 1, supplementary materials). There were clear temporal changes in mean serial interval estimates within each epidemic wave studied and across waves, with mean serial intervals shortening from 5.5 days (95% CrI: 4.4, 6.6) for pre-peak to 3.2 days (95% CrI: 1.9, 4.4) for post-peak in the second wave, and from 4.6 (95% CrI: 4.1, 5.0) days to 2.7 (95% CrI: 2.2, 3.2) days in the third wave (**Fig. 1A** and table S2). However, less clear changes were identified in the fourth wave where two peaks in incidence were noted (**Fig. 1A**). The standard deviations of serial interval distributions within individual epidemic waves were more comparable but inter-wave differences were noticeable ranging from 2.4 days to 4.3 days (table S2). The mean onset-to-isolation intervals had a similar decreasing temporal pattern along with mean serial intervals during the second wave shortening from 5.8 days to 3.5 days, while the estimates were largely constant during the third wave between 5.2 days and 5.4 days and fluctuated throughout the fourth wave (3.4 days to 5.0 days) (**Fig. 1A** and table S2). In the early part of the fifth wave, the mean serial interval was estimated to be 3.6 (95% CrI: 3.5, 3.7) days with standard deviation 3.4 (95% CrI: 3.3, 3.5) days with comparable estimates for the Delta variant (*μ*: 4.0 days, *σ*: 2.4 days), the Omicron BA.1 (*μ*: 3.3 days, *σ*: 2.0 days) and the Omicron BA.2 (*μ*: 3.6 days, *σ*: 1.8 days) subvariants.

The time-varying mean effective serial intervals and onset-to-isolation intervals across the second, third and fourth epidemic waves in Hong Kong showed similar temporal patterns in the two epidemiological parameters based on the daily changes in the estimates (**Fig. 1, B to D**). The daily variations (shortening and lengthening) in the estimates of two parameters either occurred towards the same direction (most of the second wave, some phases of the third and the fourth waves) or opposite directions (some of the phases in the third wave and the fourth wave) over the epidemics. Similar results were also indicated in the sensitivity analysis on all transmission pairs (fig. S2).

The implemented PHSMs could explain up to 70%, 42% and 49% of the variance in the mean effective serial intervals estimated for the second, third and fourth waves, respectively (table S3). The onset-to-isolation interval, reflecting the impact of the targeted measure of case identification and isolation, was found to have statistically significant positive associations with the effective serial intervals for the second and third waves with up to 60% and 13% of the variations explained in the estimated effective serial intervals for the two waves respectively, but there was not a statistically significant association in the fourth wave. The interrupted regression further suggested similar associations between onset-to-isolation intervals and effective serial intervals during different phases of the epidemics (table S4). A maximum of 10%, 29% and 48% of the variances in the estimated serial intervals were explained in the three waves respectively by both types of PHSMs (community-wide and case-based measures) when adjusting for the possible delays in the impact of onset-to-isolation. The relative population mobility and daily tests per capita showed a positive and a negative association, respectively, with the mean effective serial interval across the waves (figs. S7 to S9 and table S5).

### Factor-specific effective serial intervals

We also explored the association between the estimated serial intervals and characteristics of the infector in the identified transmission pairs in the second, third and fourth wave, including age, sex, source of infection, transmission setting, severity outcomes and onset-to-isolation delay (section 4, supplementary materials). The mean serial interval estimates ranged from 2.4 days to 5.6 days across the strata of these characteristic factors for infectors (tables S6 and S7, and fig. S3). We found that an older age (≥ 65 years) of the infectors was associated with a higher estimate of the mean serial interval (p-value <0.023) across the waves (fig. S3A and tables S7). The age-specific transmission matrices indicated that a higher proportion of the secondary infections transmitted to younger (<35 years) infectees from the infectors of all age-groups (**Fig. 2, A to C**) and initially (during pre-peak) infections started with older (>65 years) infector-infectee transmissions, then involved more younger population during peak and post-peak periods of the third and fourth waves (fig. S4). The age specific mean serial intervals were longer for older infectors (e.g., ≥ 65 years) (**Fig 2, D to F** and fig. S5).

**Fig. 2.**
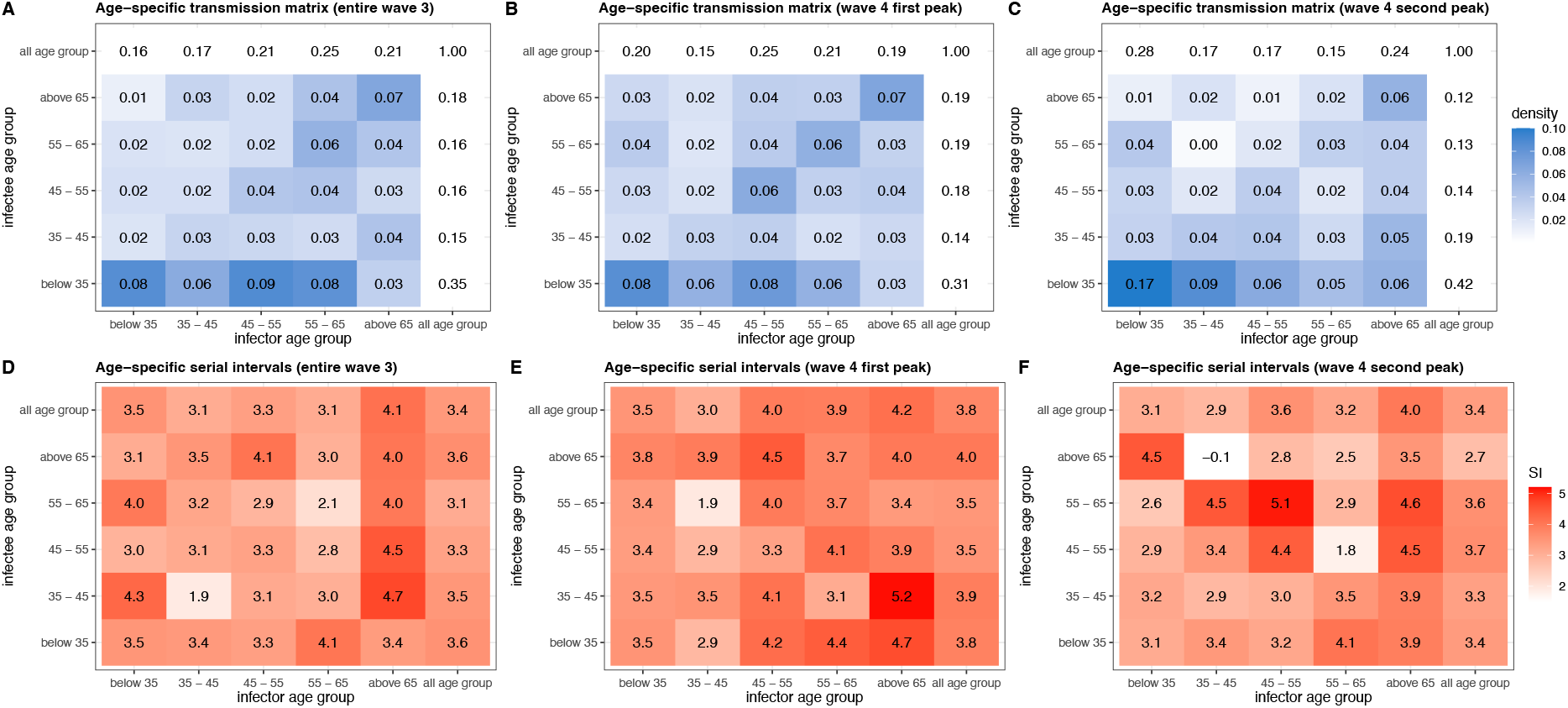
Age-specific transmission and serial interval estimated between infectors and infectees for COVID-19 in Hong Kong. The heat maps for age-specific transmission densities (the relative frequency matrices of the age distribution of infector-infectee transmission pairs for the age-groups of below 35, 35-45, 45-55, 55-65, above 65 years) with the marginal densities were for third wave (A), 1^st^ peak of fourth wave (B) and 2^nd^ peak of fourth wave (C) of COVID-19 in Hong Kong. Age-specific empirical mean serial intervals with the respective marginal estimates, evaluated for these age-groups stratifications for infector and infectee across the third wave (D), 1^st^ peak of fourth waves (E) and 2^nd^ peak of fourth wave (F). Note that no infector found of age 65 years and over during second wave, hence excluded for age-specific analysis.

In the second wave, transmission setting was identified as a statistically significant explanatory factor (p-value = 0.037) with a higher estimated mean serial interval for household transmission (tables S6 and S7). In general, the estimates of mean serial intervals were longer for the transmission pairs with critical/severe infectors across the waves, and was statistically significant for the third wave (p-value = 0.037) and fourth wave (p-value = 0.022). The onset-to-isolation intervals of the infectors were associated with estimated serial intervals (p-value < 0.005) across the waves, with shorter mean onset-to-isolation intervals correlating with shorter mean serial interval estimates. Similar results were found in the sensitivity analysis for pre-defined thresholds from (5-14) days (table S6).

### Correction in estimation of transmissibility accounting for time-varying effective serial interval distributions

We estimated time-varying *R*_*t*_ to infer changes in the transmissibility of COVID-19 in Hong Kong using the Wallinga-Teunis method (*4*) (section 5, supplementary materials) which constructed the relative likelihood for a given case being a potential infector of other cases based on the case-based reproduction number (*23*). We extended the method by incorporating the effective serial interval distribution into the estimation of *R*_*t*_ via the *EpiEstim* package in R. The estimated *R*_*t*_ from time-varying effective serial intervals was compared with that based on a constant serial interval distribution over the epidemic waves.

A simulation based modelling framework was also applied to quantify the bias from using a constant over time-varying serial interval in estimating *R*_*t*_ through a comparison of the attack rates (or cumulative number of infections) under both approaches (section 7, supplementary materials). The time series of transmission rate *β*_*t*_ derived first from the *R*_*t*_, estimated based on time-varying effective serial interval distributions and possible choices of constant serial interval distributions. Then these *β*_*t*_ were used to reconstruct the respective epidemic curves by simulating Susceptible-Infected-Recovered models to assess the related bias as the difference in attack rates from the reported data.

The differences between *R*_*t*_ estimated from real-time effective serial intervals and a constant serial interval distribution indicated that the latter might have introduced biases in inferring the transmissibility of COVID-19 over these 3 waves under analysis (**Fig. 3**). Smaller biases in *R*_*t*_ were identified during the fourth wave (except initial days) (**Fig. 3C**), during which effective serial interval distributions were relatively more stable (i.e., less varying) (**Fig. 1D**). Furthermore, with possible choices of single constant mean (*μ*) and standard deviation (*σ*) of serial interval distributions as estimated from our data, we predicted the bias in *R*_*t*_ by evaluating the mean absolute deviation from the respective *R*_*t*_, calculated by using effective serial intervals. The magnitude and direction (over-or under-estimate) of these biases depends on the mean as well as the standard deviation of the fixed serial interval distributions (**Fig. 3, A to C** and fig. S6). We found biases in simulated attack rates, were smaller when generated by considering the effective serial interval distributions over the respective choices of constant serial interval distributions (**Fig. 3, D to F** and table S8).

**Fig. 3.**
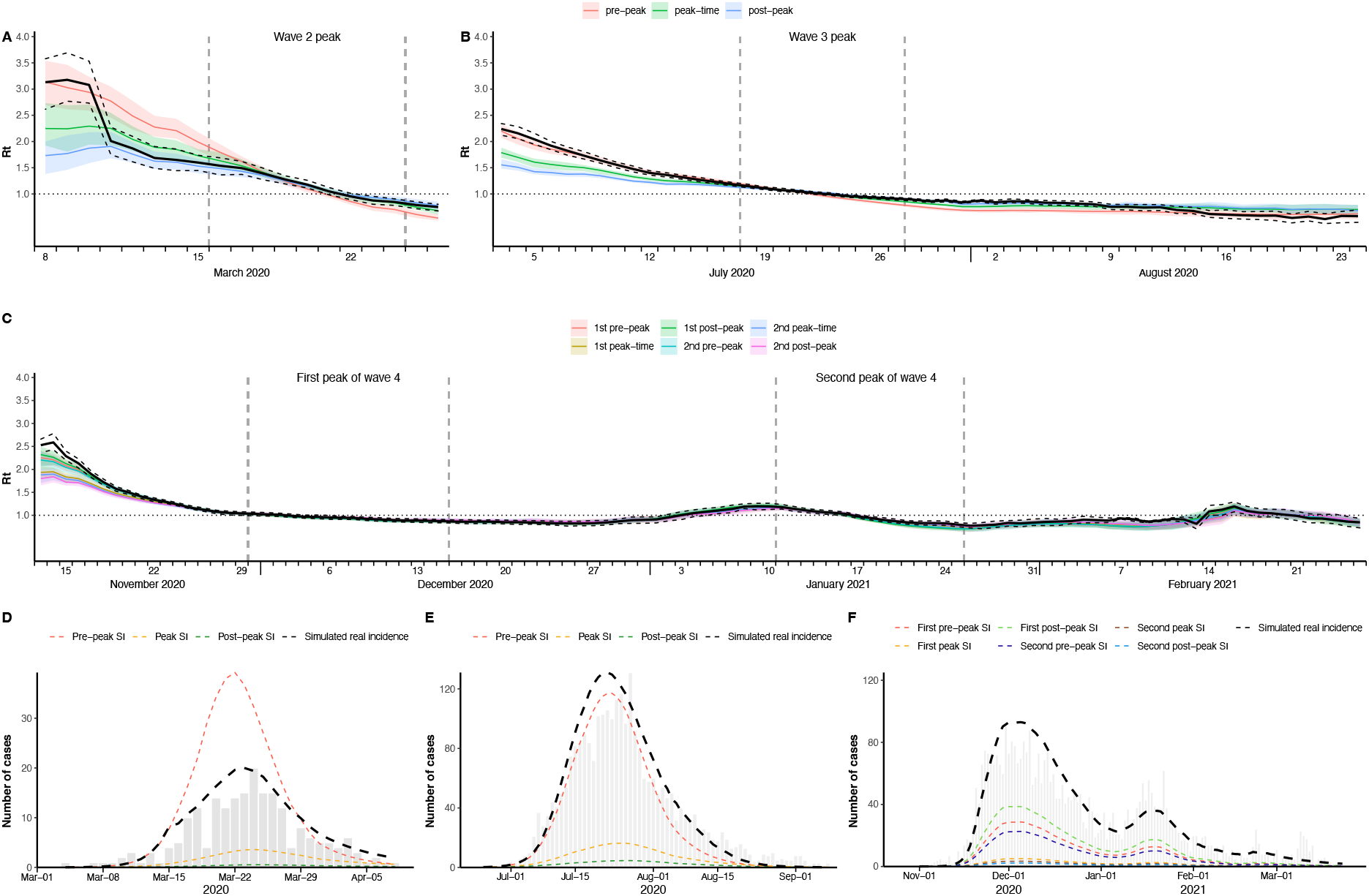
Correction in transmissibility for the bias from using constant serial interval over effective serial interval distributions. Comparison of daily estimates of effective reproduction numbers (*R*_*t*_) by using real-time effective serial interval distributions versus using a single fixed serial interval distributions for second wave (A), third wave (B) and fourth wave (C) in Hong Kong. The black solid lines are mean *R*_*t*_ (with 95% CI in dashed black lines), evaluated using effective serial interval distributions and other solid colours lines represent the mean *R*_*t*_ (with 95% CI in respective colours shades), evaluated using single fixed serial interval distributions. The fixed serial interval distributions with means (standard deviations) were considered for pre-, during and post-peak respectively in each wave as mean 5.5 (2.4) days, 4.5 (3.2) days and 3.2 (2.5) days for second wave; 4.6 (3.9) days, 3.1 (3.2) days and 2.7 (4.3) days for third wave; 4.0 (2.8) days, 3.5 (3.1) days and 4.0 (2.4) days for 1^st^ peak of fourth wave; and 4.0 (3.0) days, 3.1 (2.7) days and 3.2 (3.3) days for 2^nd^ peak of fourth wave. The differences between the black solid lines and the coloured solid lines indicate the respective biases on using single constant serial interval distribution to estimate *R*_*t*_. (D)-(E) The biases in terms of attack rates, estimated by reconstructing the epi-curves via. using the transmissibility (*R*_*t*_) derived by effective and constants serial interval distributions and Susceptible-Infected-Recovered (SIR) models for second wave (A), third wave (B) and fourth wave (C) in Hong Kong. The observed epi-curves (in grey bars) of the COVID-19 cases, the reconstructed epi-curves in dashed lines by using effective serial interval distributions (in black dashed lines) and different counterfactual constant serial interval distributions (in dashed coloured lines). The differences between the black dashed lines and the coloured dashed lines indicate the respective biases in attack rates on using constant serial interval distributions.

## Discussion

The transmission dynamics and epidemiological characteristics of COVID-19 varied significantly over time during the earlier epidemic waves studied in Hong Kong (**Fig. 1 and Fig. 3, A to C**), accounting for changes in case profiles and the impact of various control measures implemented at that time (table S1) (*16, 17, 24*). Our results for Hong Kong align with earlier findings for mainland China, in identifying a shortening in mean serial intervals over time as an epidemic is controlled with non-pharmaceutical interventions (*9, 11*). The first two waves in Hong Kong were notably driven by imported cases and potential super-spreading events (*17*), and the proportion of asymptomatic cases was found to be significantly higher during later waves (15% - 23%). The proportion of pre-symptomatic transmissions was up to 10%, and was significantly higher during the third wave, and comparable with the reported estimates elsewhere (*18, 25*). These inter- and intra-wave variations in case profiles along with varied PHSMs could shape the epidemiological parameters and hence the transmission dynamics of COVID-19 waves in Hong Kong.

In early 2020, we reported substantial reductions in mean effective serial intervals over time in mainland China following the implementation of various PHSMs including timely case isolation (*9*). Here we observed similar results in the second wave in Hong Kong (**Fig. 1B**), during which the mean onset-to-isolation delay had a positive association with mean serial interval and could explain a high proportion (59%) of variance in effective serial intervals. Other studies illustrated the effectiveness of the isolation of symptomatic cases and their potential contacts in achieving sustained control of COVID-19 transmission (*26, 27*). Strict isolation of cases will reduce transmission later in the infectious period (*9-11*). The measure of case isolation improved (i.e., early isolation) over time in Hong Kong (*28*) and appeared to be comparatively stable in third and fourth waves and have a lesser impact on the temporal changes in mean serial intervals (tables S3 and S4), which might have been associated with the broader application of PHSMs or demographic factors.

The community-wide PHSMs had negative association with serial interval across the waves, while case-based PHSMs, including case isolation had positive association with serial interval in the second and third wave (p-value = 0.33) (table S3). The association of these PHSMs with effective serial intervals could be positive or negative based on the impact of these interventions which could affect the infectors’ infectiousness profile (as truncated or modified by case-based measures including case isolation) and effective contact pattern of infectees with potential infectors (as reduced by community-wide measures and mass testing) respectively (*9-11*). This finding could suggest the lengthening of mean serial intervals during post-peak of third wave (during August 2020), when the per capita testing volume (as proxy of case-based measures) started to decline (fig. S7 and table S5), which might be attributable to missing of effective contact tracing and delays in isolation of cases (*29, 30*). The negative association between serial interval and community-wide PHSMs (table S3), could delay in finding infectees for an infector by reducing the probability of effective contacts and resulted in reverse with the strengthening of community-wide PHSMs (*30, 31*).

Along with the PHSMs, we found that certain demographic characteristics of the infectors and infectees were associated with variations in serial intervals (fig. S3A and tables S7). The older cases might be more severe and had faster and higher viral load (*32-39*), which might have been associated with shorter latent periods and incubation periods (*36, 40-46*). Therefore, the serial intervals for the transmissions from older infectors to younger infectees could be the longest or vice-versa as illustrated in fig. S10. For example, we noticed from early August 2021, during post-peak of third wave, the mean serial interval estimates started to be longer although the onset-to-isolation intervals were more or less stable (**Fig. 1C**). Although we noticed on an average the infectors were older than the infectees in the transmission pairs throughout the third wave and fourth waves (except during the end of fourth wave) (fig. S11). Temporal variation in the age distributions of infector and infectee for a transmission chain could reshape the serial interval (fig. S4). The mean infectors’ age substantially increased after the peak of third wave (from late June and early August 2020) and during the end of the fourth wave (during February, 2021) (fig. S11), might lead longer mean serial intervals during these periods (Fig. 1).

The infectors in household settings had significantly longer serial intervals for the third wave in Hong Kong (fig. S3 and table S7), accounting the proportion of older infectors in household was higher during that wave, though mean serial intervals under such settings reported not statistically significant for the first wave in mainland China (*9, 18*). For example, the household transmission was increased over time as noticed during third wave (fig. S12) and having older-to-younger transmission across the course of each wave (fig. S4) might lengthen the serial intervals during post-peak of third wave, during which mean onset-to-isolation were much stable (**Fig. 1C**).

Furthermore, the methods of estimation may affect the estimates of the serial interval distribution (*10*). Estimating temporal serial intervals via forward-looking, backward-looking or intrinsic approach by using a cohort-based framework have respective biases to be corrected (*10, 11, 47, 48*). Therefore, the estimation of reproduction number with single constant serial interval distribution can lead to bias (*9*) which can be minimised by using effective serial interval distributions instead (**Fig. 3**, fig. S6 and table S8). This bias was noticeably lower in fourth wave (**Fig. 3**) as by then the serial interval distribution had less variation across that period (**Fig. 1**).

This study provides a unique opportunity for a temporal investigation of how mean serial intervals can change over time, both shortening as well as lengthening depending on the intensity of transmission and the various case-based and community-wide PHSMs applied in Hong Kong, as well as variation in case profile (or characteristics), including demography, clinical severity, and transmission settings between and across waves. This is the first study to identify the potential factors, which could not only shorten the mean serial intervals over time, but also could lengthen the measure, therefore the resulting temporal changes in the estimates were accounted the direction and their strength of impacts. However, there are some limitations to our work. First, the change of the forward-looking serial intervals could be explained by both the PHSMs and internal mechanism (backward incubation period of infectors) (*11*), but detailed exposure information was not always available. Second, information on serial intervals collected through contact tracing could suffer various biases such as recall biases, which could reduce the chance of identifying longer serial intervals for example. However, our intense algorithmic check and cross check with additional phylogenetic data should minimise the impact of this biases on the main outcomes. Finally, while we identified the potential for bias in *R*_*t*_ estimates if using constant serial interval distributions, there are other potential biases in *R*_*t*_ estimation that we did not discuss here.

In conclusion, our results indicated that the changes in serial interval distributions might not be always monotonic as reported earlier for mainland China in 2020 (*9*). The real-time variations in serial interval distributions were also driven by the changes in transmission pattern (who infects whom), characterised by temporal variation in demographical and clinical profiles of the cases along with PHSMs during the first two years of the COVID-19 pandemic in Hong Kong. This time varying matric of effective serial interval distributions could improve the estimation of transmissibility, the time varying reproduction number (*R*_*t*_) accounting for intermediate factors of transmission and allow us to assess the timely impact of public health measures.

## Data Availability

All data, code, and materials used in the analyses in main text and supplementary materials will be available together with the publication of this paper.

## Acknowledgments

We acknowledge Ms. Julie Au for technical assistance. We thank the Centre for Health Protection in Hong Kong for their data collection and compilation throughout the COVID-19 pandemic. We thank Kathy Leung for helpful discussions. STA, DC, and acknowledge the research computing facilities and advisory services offered by Information Technology Services, The University of Hong Kong.

## Funding

Health and Medical Research Fund, Health Bureau, Government of the Hong Kong Special Administrative Region grant COVID190118 (BJC) and grant 20190712 (STA);

Collaborative Research Scheme grant C7123-20G and grant T11-705/14N, Research Grants Council, Government of the Hong Kong Special Administrative Region (BJC);

AIR@innoHK program of the Innovation and Technology Commission, Government of the Hong Kong Special Administrative Region (STA, ZD, EHYL, PW, GML, BJC);

European Research Council (grant no. 804744); the Grand Challenges ICODA pilot initiative, delivered by Health Data Research UK and funded by the Bill & Melinda Gates Foundation and the Minderoo Foundation (LW);

The funding bodies had no role in study design, data collection and analysis, preparation of the manuscript, or the decision to publish.

## Author contributions

Conceptualization: STA, PW, BJC

Methodology: STA, DC, LW, EHYL, BJC

Investigation: STA, DC, WWL, AY, DCA, YCL

Data: WWL, AY, DC, DCA, JX, FH, HG, STA

Visualization: DC, AY, YCL, STA

Comments: DCA, EHYL, LW, ZD, JYW, XKX, PW, GML, BJC

Funding acquisition: STA, EHYL, ZD, LW, PW, GML, BJC

Project administration: STA, DC, WWL, DCA, YCL

Supervision: GML, BJC

Writing – original draft: STA, DC, BJC

Writing – review & editing: STA, DCA, EHYL, LW, ZD, JYW, XKX, PW, GML, BJC

## Supplementary Materials

Materials and Methods

Supplementary Text

Figs. S1 to S12

Tables S1 to S8

References (*49*–*55*)

