## Supplementary Information for "Insights into COVID-19 epidemiology and control from temporal changes in serial interval distributions in Hong Kong"

**This PDF file includes:**

Materials and Methods
Supplementary Text
Figs. S1 to S12
Tables S1 to S8

**Table of Contents** **Pages**

| | | |
|---|---|---|
| **1.** | **Contact tracing data and transmission pairs constriction** | **3** |
| **2.** | **Estimation of serial interval and onset-to-isolation interval distributions** | **6** |
| **3.** | **Multivariable Regression Analyses** | **7** |
| **4.** | **Stratification of the transmission pairs and assessment of the factors for serial intervals** | **9** |
| **5.** | **Estimation of instantaneous reproduction number (Rt)** | **10** |
| **6.** | **Sensitivity Analyses** | **11** |
| **7.** | **Simulation framework** | **12** |
| **8.** | **Supplementary Figures S1-S12** | **14** |
| **9.** | **Supplementary Tables S1- S8** | **26** |
| **10.** | **References** | **37** |

### Materials and Methods

#### 1. Contact tracing data and transmission pairs construction

The contact tracing information is retrieved from the line-list data collected from the Centre for Health Protection, Hong Kong SAR government, which includes the information on confirmation/report date, illness onset date, isolation date, admission date, severity outcome (critical/serious/stable), infection origin (imported/local) and arrival date (if imported), location settings (building address and workplace), cluster settings and specific travel/movement history for each recorded confirmed case in Hong Kong (*12*).

We followed our earlier work (*17*) to reconstruct the initial pairs with reference to the COVID-19 line-list information of all cases provided by the Centre for Health Protection. We rechecked the initially constructed pairs by their cluster settings and epidemiological linkage with other cases of intra- or inter clusters and determined the infector and infectee within a pair according to a pre-defined algorithm. We checked the version of the line-list updated until 31 July 2022.

Our algorithm set the following criteria for determining which case is the infector within a larger cluster and/or smaller cluster setting, in descending order of importance:

I. Recent travel history

II. Association with larger outbreaks

III. Movement/Jobs in areas with high chances of contact with the virus (involving direct/indirect contact with the virus)

The remaining case(s) would be regarded as infectees.

Clusters consisting of 2 cases with a clear infector and infectee were deemed ‘certain’ or ‘confirmed transmission’ pairs, while two major scenarios during the rechecking led to classification of the pairs as ‘uncertain’ or ‘likely’. The first is the scenario of multiple infectors. In clusters involving more than 2 cases, the exact transmission dynamics were not clear. For example, in a cluster consisting of cases A, B and C, even if we know that A was the primary case in this cluster who introduced the virus, we do not know if B and C were both directly infected by A (A → B and C), or indirectly infected (A → B → C or A → C → B). We simplified this by treating the primary case as the infector of all subsequent cases (A → B and C) when the serial intervals between primary case and subsequent cases fall within our given threshold, details are in section 3 below.

The second scenario was that of unclear infectors. In some clusters that have complicated epidemiological linkages between cases, none of the cases could be considered an infector according to our algorithm. We tried to cross-check unclear pairs with the phylogenetic database to identify the infector-infectee relationship by using *Phybreak* package in R (*19*), but were only able to resolve 7 pairs, as genetic information was only available for 11 unclear pairs. For the remaining unclear infectors, when serial interval was within the threshold, we defined the infector as the case with the earliest onset date. If cases shared the same onset date, the report date, followed by case number were used (*9, 18, 49*) . For example, if A and B were both regarded as possible infector for C, choose the infector with earliest onset date; if A and B had same onset dates, choose the infector with earliest report date; if A and B also had same report dates, choose the infector with smaller case number in the line list, which also indicates that case was reported earlier.

Other reasons for flagging pairs as uncertain included possible separate infection, where we thought that both cases had equally high risk of being an infector according to our algorithm, and possible co-infection, where we thought that both cases could have been infected by an external common source, such as fomites. We expressed the relationship between cases in the same way as we did with the unclear infector scenario.

We did not analyse the few transmission pairs in 'first wave' during January-February, 2020, due to little local incidences. Here we define our 'second wave' of local transmission from March 1, 2020 to April 10, 2020. During this period, 911 cases were onset or reported, among whom 569 (62.5%) cases were classified as imported cases. In the remaining 342 local cases, 42 (12.3%) cases were asymptomatic. We identified 64 infectors in this period, 34 of them were onset from March 16 to March 24, therefore we defined March 16 to March 24 as the peak timing of second wave. There were 47 clearly confirmed infector-infectee pairs in second wave, and 40 more pairs have met our serial interval threshold (5-14 days) for main analysis, in total 87 pairs were used for estimating the serial interval in this period.

We define our 'third wave' from June 25, 2020 to September 8, 2020. During this period, 3728 cases were onset or reported, among whom 422 cases were classified as imported cases, while 3306 (88.7%) cases were local cases, and in these local cases 562 (17.0%) cases were asymptomatic. We identified 736 infectors in this period, 274 of them were onset from July 18 to July 27, which was more than one third of 736, therefore we defined July 18 to July 27 as the peak timing of third wave. There were 357 clearly confirmed infector-infectee pairs in third wave, and 608 more pairs have met our threshold, in total 965 pairs were used for estimating the serial interval in this period.

We define the ‘fourth wave’ from November 1, 2020 to March 23, 2021. During this period, 6089 cases were onset or reported, among whom 673 cases were classified as imported cases, while 5416 (88.9%) cases were local cases, and in these local cases 1396 (25.8%) cases were asymptomatic. We identified 1131 infectors in this period. Because the duration of fourth wave was relative longer comparing with previous periods, we defined two peaks of infection waves in fourth wave. The first peak was from November 30, 2020 to December 15, 2020, during which time there were 333 infectors onset. The second peak was from January 11, 2021 to January 25, 2021, during which time there were 191 infectors onset. There were 355 clearly confirmed infector-infectee pairs in fourth wave, and 1026 more pairs have met our threshold, in total 1381 pairs were used for estimating the serial interval in this period. For ‘fifth wave’, in early January 2022, detailed data on confirmed transmission pairs were only available in the very early stage of this wave. We identified additional total of 229 pairs (30 for BA1, 174 for BA2 and 25 for delta) identified until mid-February 2022, and not allowed us to perform temporal analyses for fifth wave (fig. S1).

### 2. Estimation of serial interval and onset-to-isolation interval distributions

We estimated the mean and standard deviation of the serial interval distribution by fitting normal distribution on the empirical data accounting for the potential of pre-symptomatic transmission (*9*). The fitting was performed in a Bayesian framework using Markov Chain Monte Carlo implemented with the *RStan* package in R. The initial values of the parameters were generated from uniform random samples, we set 1000 warm-up samples, 4000 iterations and run 4 parallel chains.

We first examined potential changes in the serial interval distribution over time in each epidemic wave, which may occur as a consequence of PHSMs or for other reasons (*9, 50*). We first estimated the serial interval distribution for inter-wave as well as before, during and after the peak in incidence in each wave, and then estimated the time-varying effective serial interval distribution with a sliding window of 10 days (*9*). We varied the sliding window from 7 days to 14 days as a sensitivity analysis (section 4 below). We estimated the mean onset-to-isolation interval (case isolation delay) of the infectors by fitting shifted gamma distributions to the empirical interval data of 10-day sliding windows using same inferential framework.

#### 3. Multivariable Regression Analyses

We performed multivariable regression analysis on mean serial intervals and mean onset-to-isolation interval as an indicator of timely case isolation, where various PHSMs were included as factorized explanatory variables. While the timing of these interventions was considered to fall on a specific window (of 10 days) for which effective serial intervals was calculated if at least $5^{th}$ day of the window contain the start or end date of a critical PHSMs, and then assigned respective PHSM level to that window. We conducted on the time window's data that are in line with presented in Fig 1 and S2, to ensure the SI values was estimated given enough sample size. For $2^{nd}$ wave, regression was conducted on data in the sliding windows from Mar 04 – Mar13 to Mar 22- Mar 31, during this time, the most critical PHSMs are testing asymptomatic inbound travelers since March 23 and suspending international airport service since March 24, thus we defined Mar 20 – Mar 29 as the start time window of PHSM level 2 in second wave.

For 3rd wave, regression was conducted on data in the sliding windows from Jun 25 – Jul 04 to Aug 20 – Aug 29, during this time, July 15 was the implementation date of group gathering no more than 4; July 29 was the implementation date of further restricted gathering limit of no more than 2 and food service suspended from 6pm to 5am. Thus, sliding window from Jul 11 – Jul 20 to Jul 24 – Aug 02 were labeled as PHSM level 2 period, Jul 25 – Aug 3 to Aug 20 – Aug 29 were labeled as PHSM level 3 period.

For 4th wave, regression was conducted on data in the sliding windows from Nov 27 – Nov 16, 2020 to Feb 22 – Mar 03, 2021, during this time there were many different PHSMs implemented, that we could distinguish them into case-based and community-based PHSM. For case-based PHSM, the most critical time periods were from Dec 02, 2020 to Feb 17, 2021, group gathering was again limited to no more than 2, so sliding window from Nov 28 – Dec 07 to Feb 13 – Feb 22 were labeled as PHSM community level 2, while other time windows were PHSM community level 1. For case-based PHSM, the first critical day was on Nov 24 when mandatory virus testing policy was extended to many risky occupation groups, while on Jan 26, 2021, restricted block lockdown and mandatory testing were implemented for risky areas. Therefore, we labeled time window from Nov 20 – Nov 29 to Jan 21 – Jan 30 as PHSM case level 2, Jan 22 – Jan 31 to Feb 22 – Mar 03 as PHSM case level 3.

The variation in population mobility and physical mixing, which also potentially reflect the impact PHSMs and might have significant impact on disease transmission of COVID-19 (*20-22*). Therefore, we additionally retrieved the digital transactions made on Octopus cards, generally used for daily public transport and small retail payments in Hong Kong from the Octopus website (https://www.octopus.com.hk/tc/consumer/index.html). We defined the daily relative mobility as the relative digital transactions made on Octopus cards with respect to the transaction on 1st January, 2020 (*20*). We considered the time series of four category of Octopus users 'Children', 'student', 'adults' and 'elderly' for travel (fig. S8) and retail (fig. S9) related uses, to identify any significant changes in these stratifications. We also retrieve the temporal (daily) testing volume in Hong Kong across the waves as reported by Department of Health, the Government of the Hong Kong SAR. We defined the per capita testing volume as daily number of tests per 10,000 population. We then performed regression analyses on these proxy time series to quantify their association with effective serial intervals (figs. S7 to S9, and table S5). Note that the time series of daily relative mobility is a proxy of community-wide PHSMs as reverse metric, where higher relative mobility proxy indicates lower impact of community-wide PHSMs (*51*). Similarly, the per capita testing volume is a reverse metric of the proxy for isolation, lower per capita testing indicates possibly lower contact tracing and delay in isolation of infectors (*52*).

**4. Stratification of the transmission pairs and assessment of the factors for serial intervals**

We stratified our transmission pair data to identify the potential factors of serial interval by infector's characteristics including: age, sex, source of infection, transmission setting, severity outcome and onset-to-isolation delay. We stratified age of infectors into two groups, below 65 and above 65 to examine the transmission dynamics between older adult over younger. Where, household and non-household transmission events were considered as the transmission settings. Severity outcome reflected the infector's clinical severity status into three levels, stable, serious and critical. The measure onset-to-isolation was stratified into shorter and longer with the reference to the median values for respective waves.

We then performed non-parametric tests (Kruskal–Wallis H test) to identify statistically significant differences in serial intervals between groups. We tested sensitivity of our main analysis by applying the stratification analysis on the datasets based on different thresholds for the onset interval between the infector and the infectee, ranging from 5 days to 14 days (section 6 below). We further explored the age specific transmission matrix to identify the age distribution of infectors and infectee in the transmission chains and estimated age-specific serial intervals and compare them for inter and intra-waves (pre-, during and post-peaks). We estimated the mean serial interval and mean onset-to-isolation interval (with respective uncertainty) for the age-groups (below 35, 35-45, 45-55, 55-65, and above 65 years) of infector-infectee transmission pairs during pre-peak, peak, and post-peaks timing. We further estimated the serial intervals for different variants in Hong Kong including wild-variant for up to fourth wave and delta, omicron AB1, AB2 variants for initial fifth wave.

### 5. Estimation of effective reproduction number ($R_t$)

Our estimation of $R_t$ was based on Wallinga & Teunis method, (*4*) and implemented through *wallinga_teunis* function from *EpiEstim* package in R. $R_t$ is defined as the average number of secondary cases that were infected by one primary case with onset at time $t$. The Wallinga & Teunis method provides a likelihood-based estimate of effective reproduction number, accounting the infector's perspective, which is basically the temporal order of cases reproduction number (*23*).

Consider, case $i$ has been infected by case $j$ with their time of symptom onset at $t_i$ and $t_j$ respectively. Therefore, the relative likelihood $P_{ij}$ can be expressed in terms of the probability distribution for the generation interval $w(\tau)$ as,

$$P_{ij} = \frac{w(t_i - t_j)}{\sum_{i \neq k} w(t_i - t_k)}$$

Then the case reproduction number for case $j$ is sum of the relative likelihood of the case $i$ being infector of all other cases in given cohort. Therefore, case reproduction number, $R_j = \sum_i P_{ij}$ and then simply ordering these case $R_j$ by their timing of infection (proxied by timing of symptom onset), we would get the effective reproduction number $R_t$ for a given period $t$.

We further developed the method simply incorporating the effective serial interval distribution via *EpiEstim* package. We allowed 100 sets of simulation for estimating confidence interval. To incorporate with our temporal effective serial intervals, we embedded the length of sliding window in *wallinga_teunis* function as 10 days (same length of sliding window used for effective serial interval estimation) and ensured the start and end of the sliding windows for $R_t$ should correspond to the sliding windows used for effective serial interval. We also explored the real-time measure of transmissibility as instantaneous reproduction number proposed by Cori et al (*3*) but found not reasonable as our effective serial interval distribution are forward looking of a given cohort. Whereas the effective reproduction number $R_t$, proposed by Wallinga & Teunis (*4*), which is a forward measure and in the same line of the estimation of the effective serial interval distribution.

### 6. Sensitivity Analyses

A significant number of pairs were uncertain pairs. We considered a range of possible serial intervals accounting household and non-household transmission settings, to have a reliable threshold for resolving the uncertainty. We first estimated mean ($\mu$) and standard deviation ($\sigma$) of the serial interval of the confirmed pairs first and found was 3.92 (95% CrI: 3.60, 4.24) and 4.55 (95% CrI: 4.34, 4.79) respectively. Therefore, we set the upper 1 $\sigma$-limit ($\mu + \sigma$), i.e., approximately 8 days as the threshold to include the uncertain pairs with a maximum serial interval of 8 days for our main analysis. A sensitivity analysis has been performed with different choice of the threshold from 5 day to 14 days. We found the choice of 8 days was consistent in showing the different patterns of stratified estimation and inter-wave changes, while also contained less variance in the estimates.

Along with the above data level sensitivity analysis, we performed some sensitivity analyses at method level including sensitivity of sliding windows for estimation effective serial intervals. We allowed 7 days to 14 days of window lengths. We used the estimates in 10-day sliding window as our main analysis, because the choice of 7 days would have lower sample size in the beginning windows, while the choice of 14 days would provide too smoothed estimates and could not clearly capture the temporal changes when the length of the total period was short.

### 7. Simulation framework

We used Susceptible-Infected-Recovered (SIR) models to reconstruct the observed epi-curves by evaluating the time series of transmission rate $\beta_t$ from these $R_t$ (*53, 54*), estimated by time varying effective serial intervals and various choices of constant serial intervals. For simplicity, we considered constant serial interval distribution estimated for the data during pre-peak, peak timing and post peak of each wave. Note that the fourth wave had two peaks, therefore had 6 choices of such constant serial interval distributions to consider. We analysed and compared (by evaluating the attack rates/ cumulative number of infections) these epidemic curves with the fitted epi-curves using predicted effective serial intervals for each waves in Hong Kong. We considered no prior immunity (for simplicity) and set the initial conditions for these simulation as I(0) in the range

[0.00001, 0.00005], recovery rate ($\gamma$) set to the range [0.20, 0.67] i.e., mean infectious period up to 5 days across the wave (*55*).

### Supplementary Figures

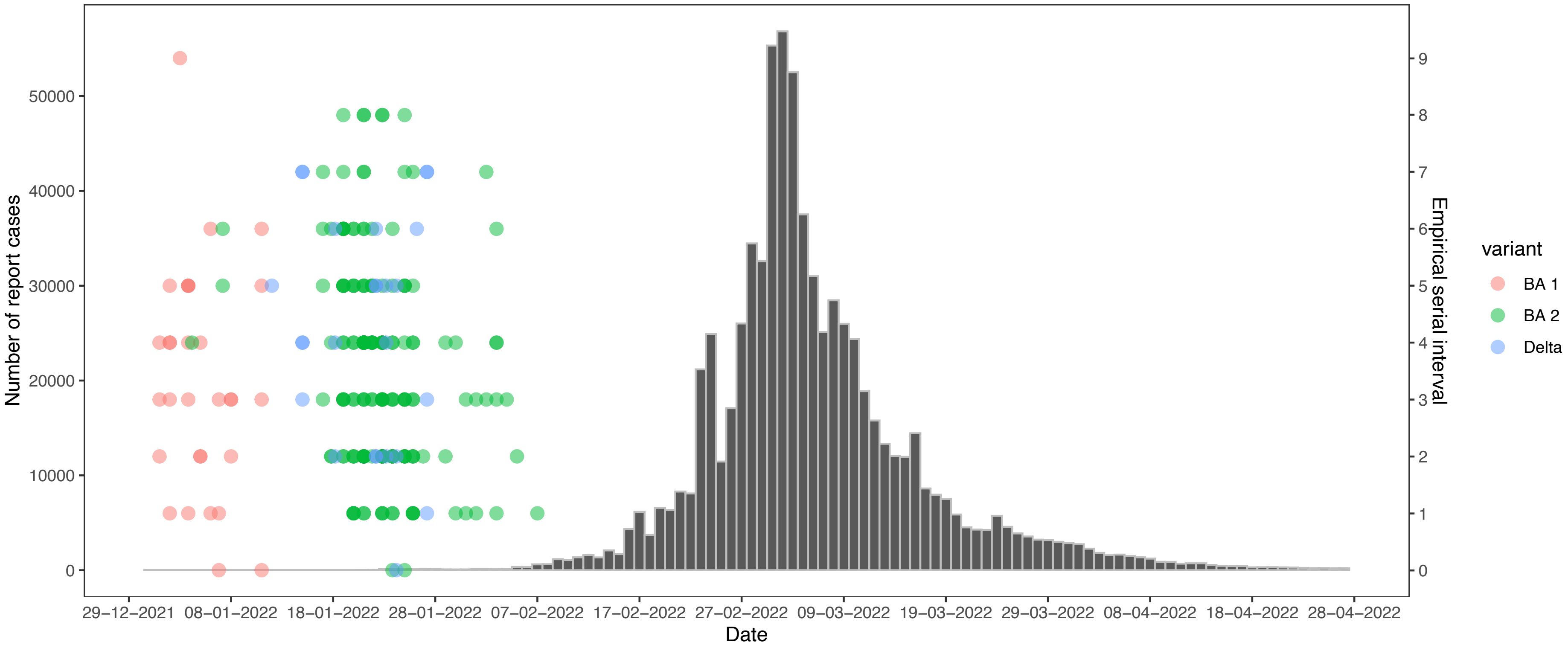

**Fig. S1. Transmission dynamics and empirical mean serial intervals for fifth-wave of COVID-19 in Hong Kong.** The grey bars indicate the epi-curve of the reported COVID-19 cases during fifth wave in Hong Kong. Empirical mean serial interval estimates for different variants for delta and omicron (BA1 and BA2) are in colored dots during early fifth wave (until mid-February 2022) in Hong Kong. The information on detailed data for confirmed transmission pairs were not available after mid-February, 2022 for this wave.

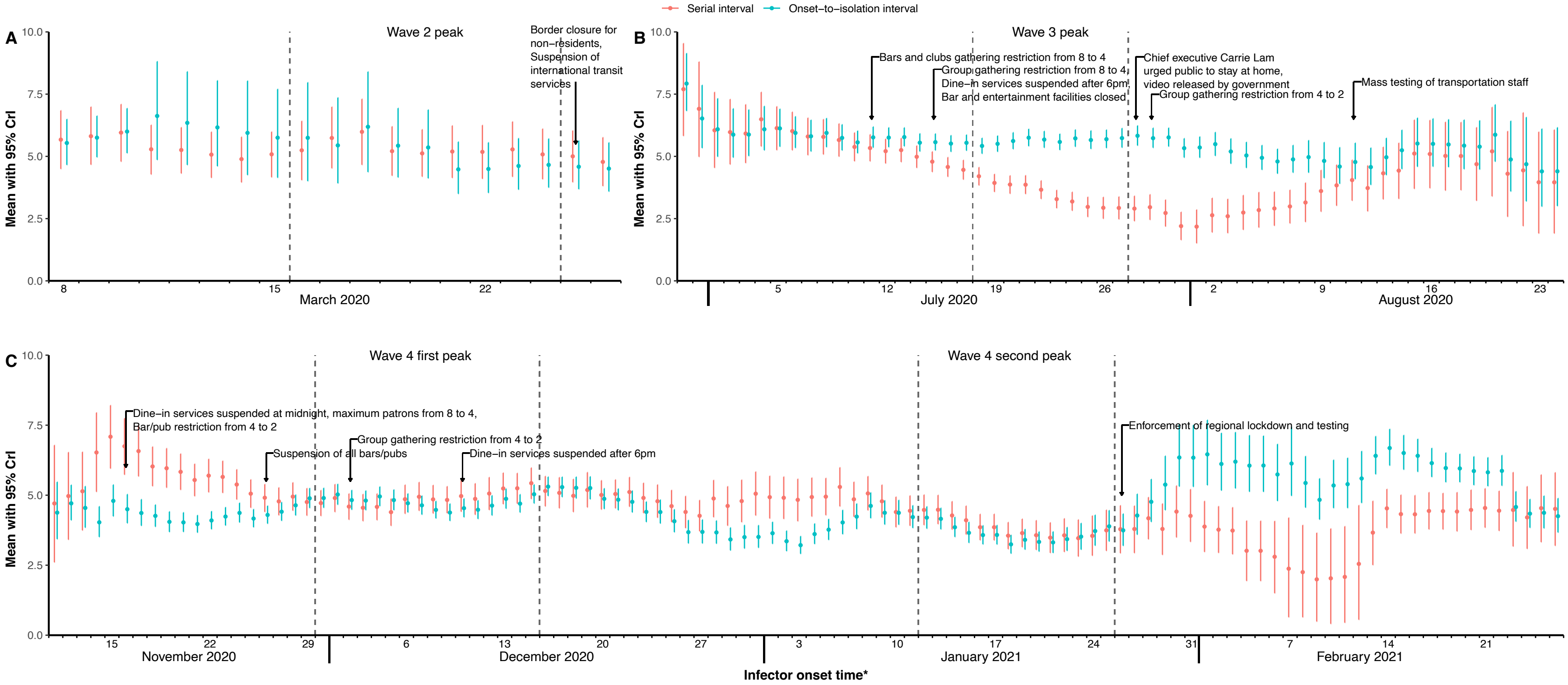

**Fig. S2. Temporal estimates of serial interval distribution and isolation delay (for all transmission pairs) for COVID-19 in Hong Kong**. Time varying estimates of effective serial intervals and onset-to-isolation interval, evaluated from all confirmed and likely transmission pairs (irrespective of any predefined thresholds) for second (A), third (B) and fourth (C) waves with the indicator to timings of some significant public health and social measures (PHSMs) in Hong Kong. The estimates presented here are based on 10-day sliding windows evaluated via MCMC as mean effective serial intervals (red dots) with 95% CrI (in red vertical line segments) and mean onset-to-isolation interval (in teal dots) with 95% CrI (in teal vertical line segments). The effective serial intervals and onset-to-isolation interval were obtained by fitting the empirical data to the normal distribution and gamma distribution respectively. The grey dashed regions indicate the peak timing of each wave (presented with respect to the 5th day of each sliding window). The estimates are presented for the days with sufficient transmission pairs ($\geq 20$) in each sliding window to ensure the stable estimates during the wave ends particularly for 3$^{rd}$ and 4$^{th}$ waves.

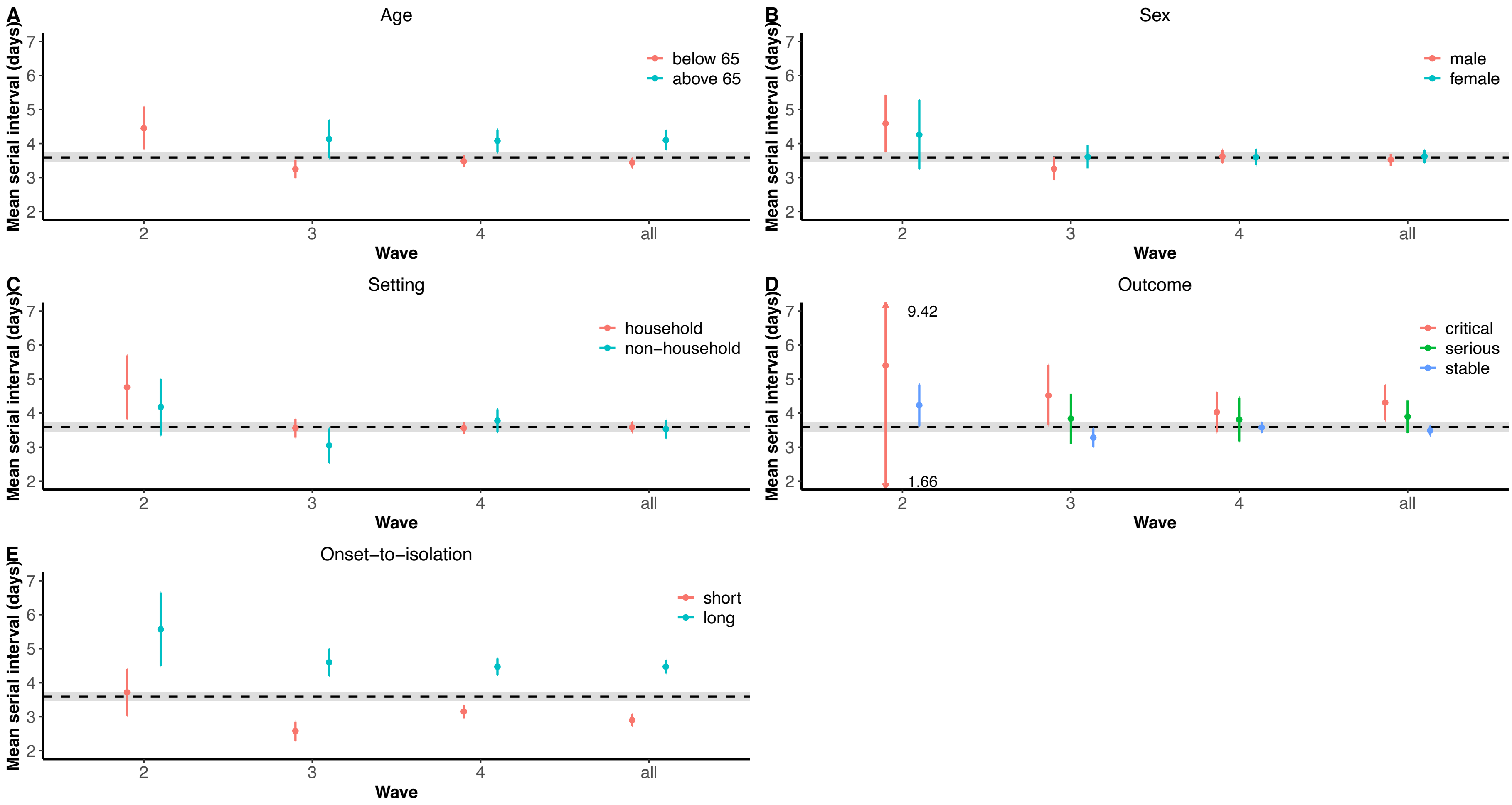

**Fig. S3. Factor specific estimates of serial intervals of COVID-19 in Hong Kong.** The mean serial interval estimates (in dots) with 95% CrI (in the vertical line segments) evaluated by fitting normal distribution using MCMC for various factor stratifications including infector age (A), infector sex (B), transmission setting (C), infector severity outcome (D) and infector onset-to-isolation interval relative (E). The black horizontal dashed lines indicate the overall (without any stratifications) mean serial interval 3.59 days with 95% CrI (3.46, 3.73) days (in grey shaded region).

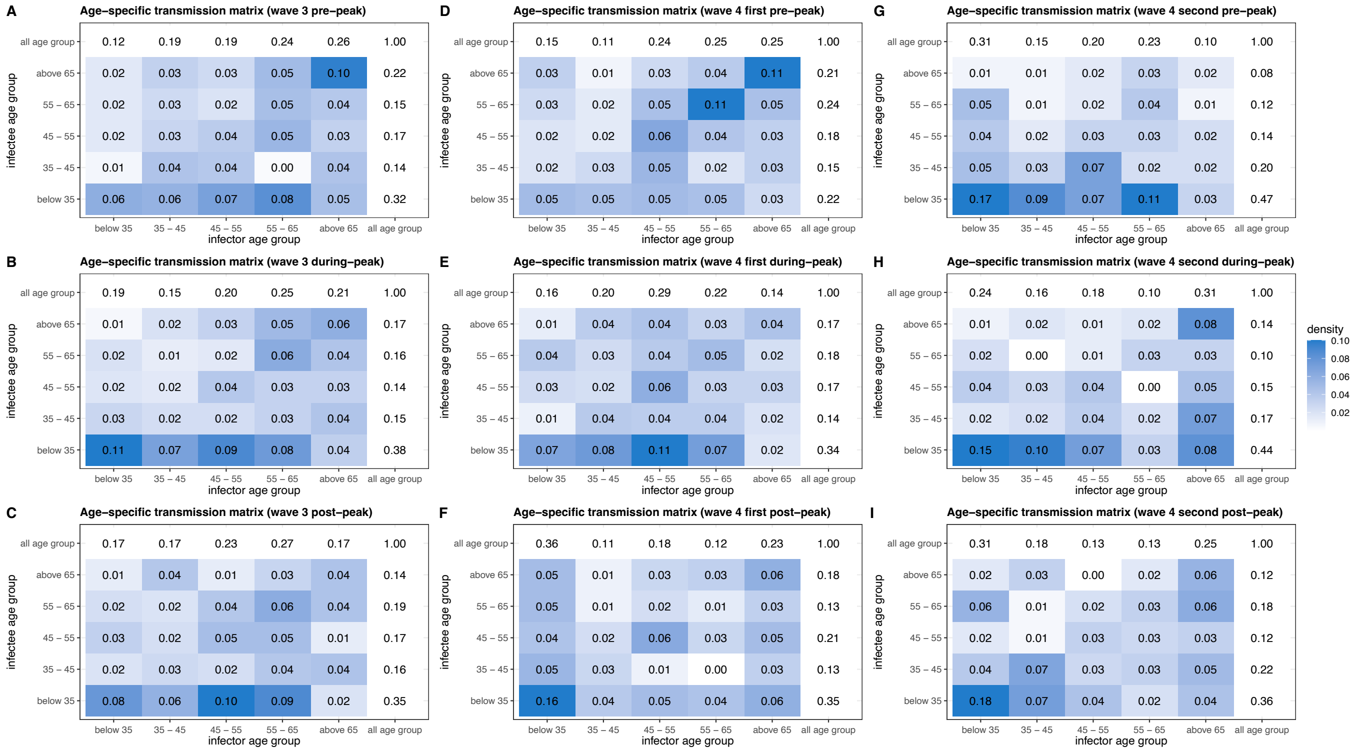

**Fig. S4. Age-specific and temporal (pre-, during and post-peak) transmission for third and fourth wave of COVID-19 in Hong Kong.** The heat maps for age-specific transmission densities (the relative frequency matrices of the age distribution of infector-infectee transmission pairs for the age-groups of below 35, 35-45, 45-55, 55-65, above 65 years) with the marginal densities, calculated for during pre-peak, peak and post-peak timing for third wave (A-C), 1st peak of fourth waves (D-F) and 2nd peak of fourth wave (G-I).

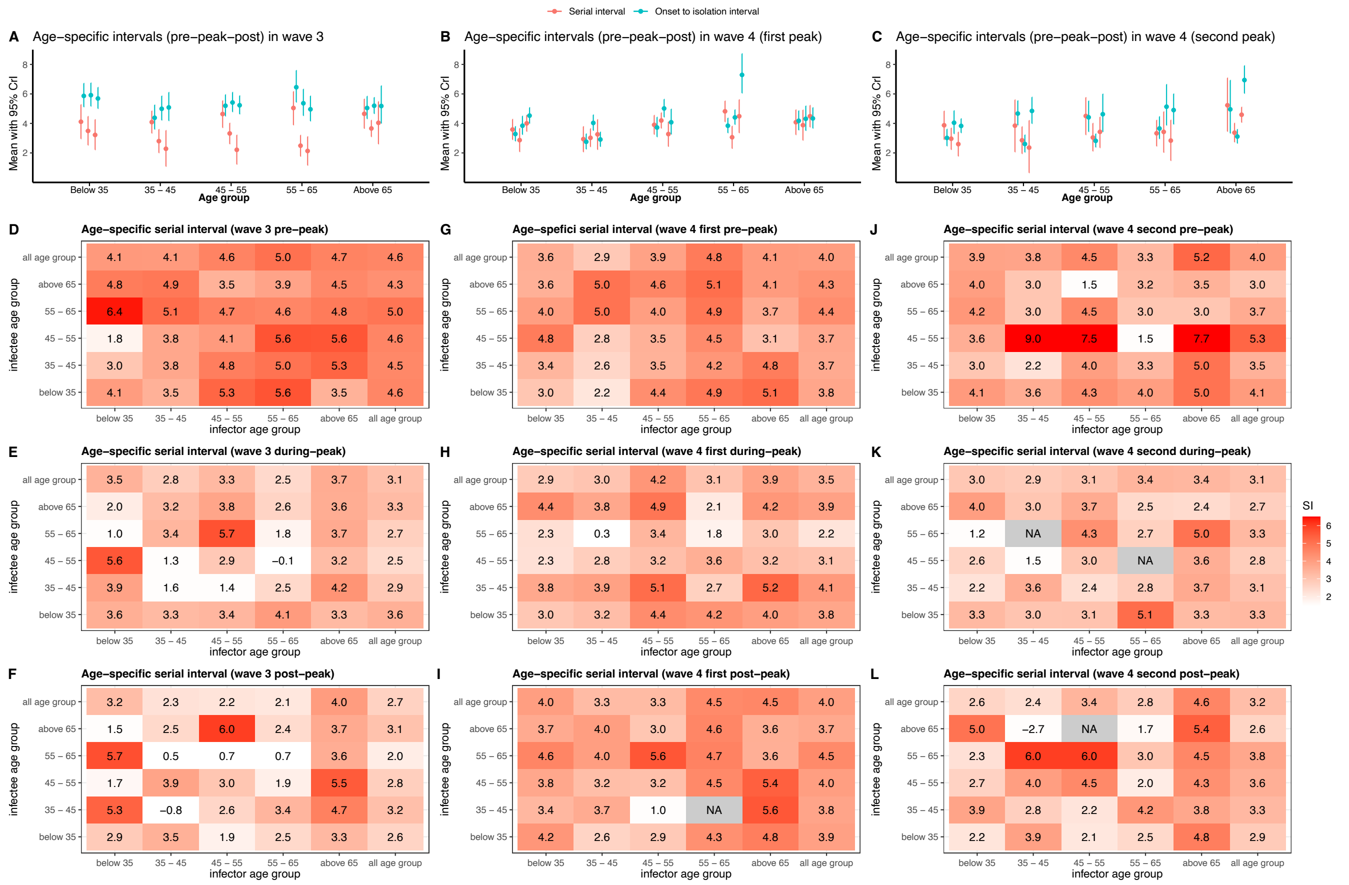

**Fig. S5. Age-specific and temporal (pre-, during and post-peak) mean serial intervals and onset-to-isolation intervals for third and fourth wave of COVID-19 in Hong Kong.** (A)-(C) Mean serial interval estimates (red dots) with 95% CrI (in red vertical line segments) and mean onset-to-isolation interval (in teal dots) with 95% CrI (in teal vertical line segments), evaluated for the age-groups (below 35, 35-45, 45-55, 55-65, above 65 years) of infectors in transmission pairs during pre-peak, peak, and post-peaks timing for third wave (A), 1st peak of fourth waves (B) and 2nd peak of fourth wave (C) respectively. (D)-(L) The heat maps for age-specific empirical mean serial intervals with the respective marginal estimates, evaluated for these age-groups stratifications of infector and infectee across the third wave (D-F), 1st peak of fourth waves (G-I) and 2nd peak of fourth wave (J-L).

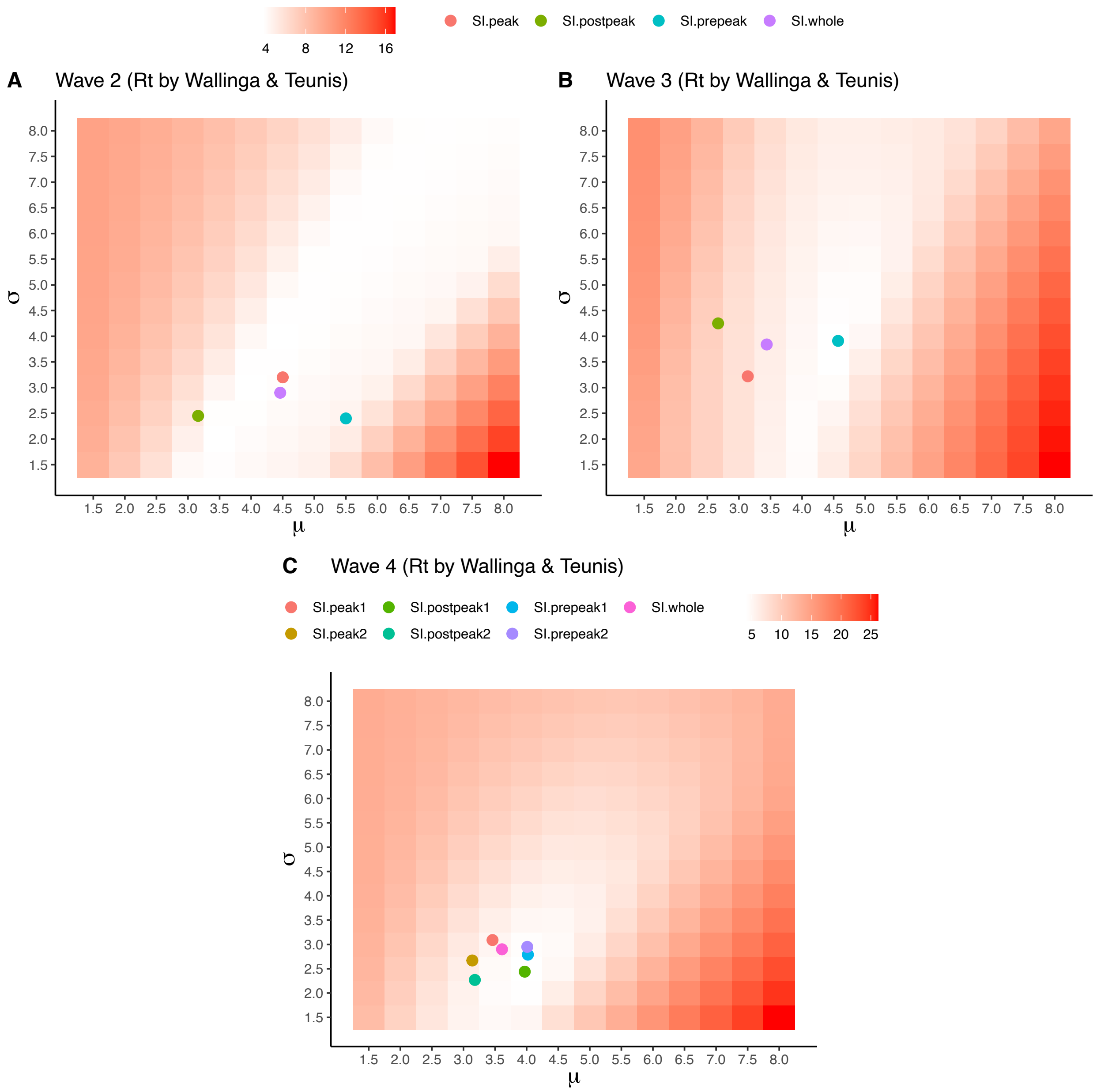

**Fig. S6. The heatmaps to present the biases in estimating $R_t$ using single constant serial interval distribution with different choices of the mean ($\mu$) and standard deviation ($\sigma$) over time-varying serial intervals during the epidemic.** The color gradient indicates the value of the biases as the mean absolute deviation of $R_t$ estimated by comparing these choice of serial interval distributions and the effective serial intervals for second (A), third (B) and fourth (C) wave respectively. The bold points in different colours indicate such choice of single serial interval distributions and their related biases.

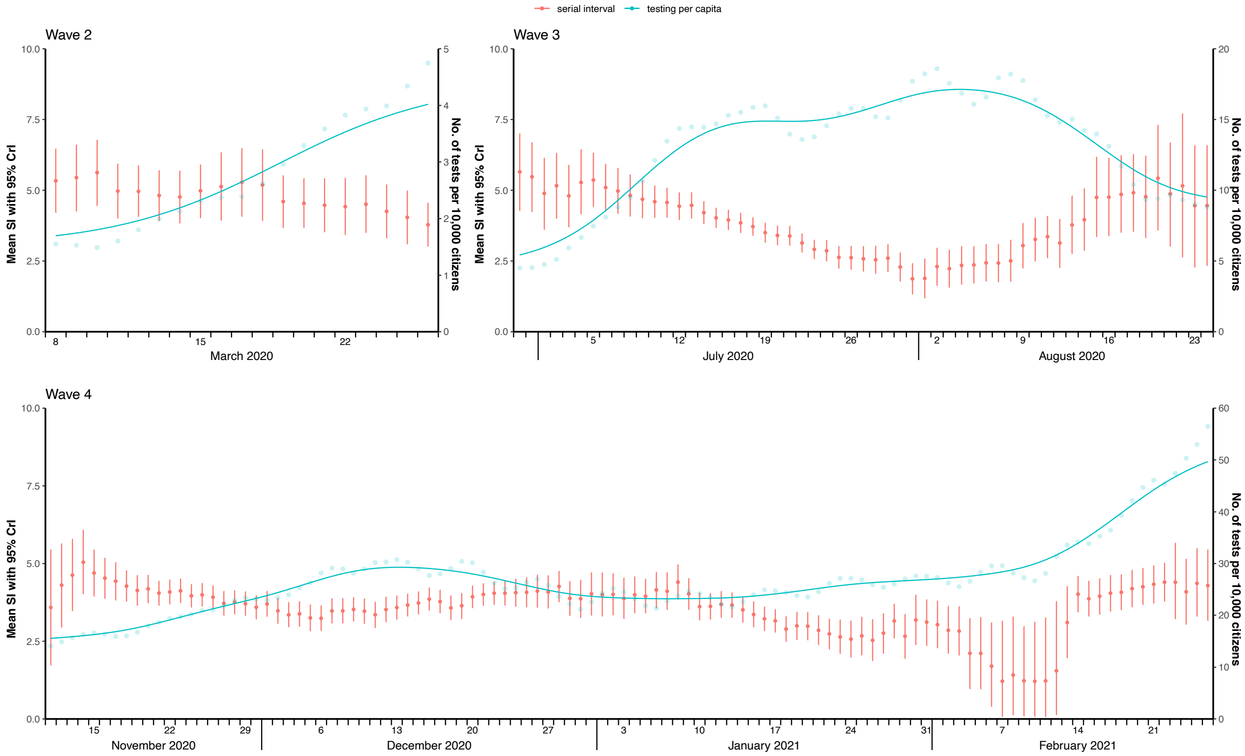

**Fig. S7. Effective serial interval estimates, time series COVID-19 testing volume per capita across the waves of pandemic in Hong Kong.** The time varying estimates of effective serial intervals in black dots with 95% CrI (in black vertical line segments). The daily number of tests per 10,000 population presented in the blue dots for 2$^{nd}$ and 3$^{rd}$ waves with their fitted lines (non-linear kernel regression) in blue lines, calculated based on the empirical data in 10-day sliding windows (presented with respect to mid-date as the 5th day of each sliding window).

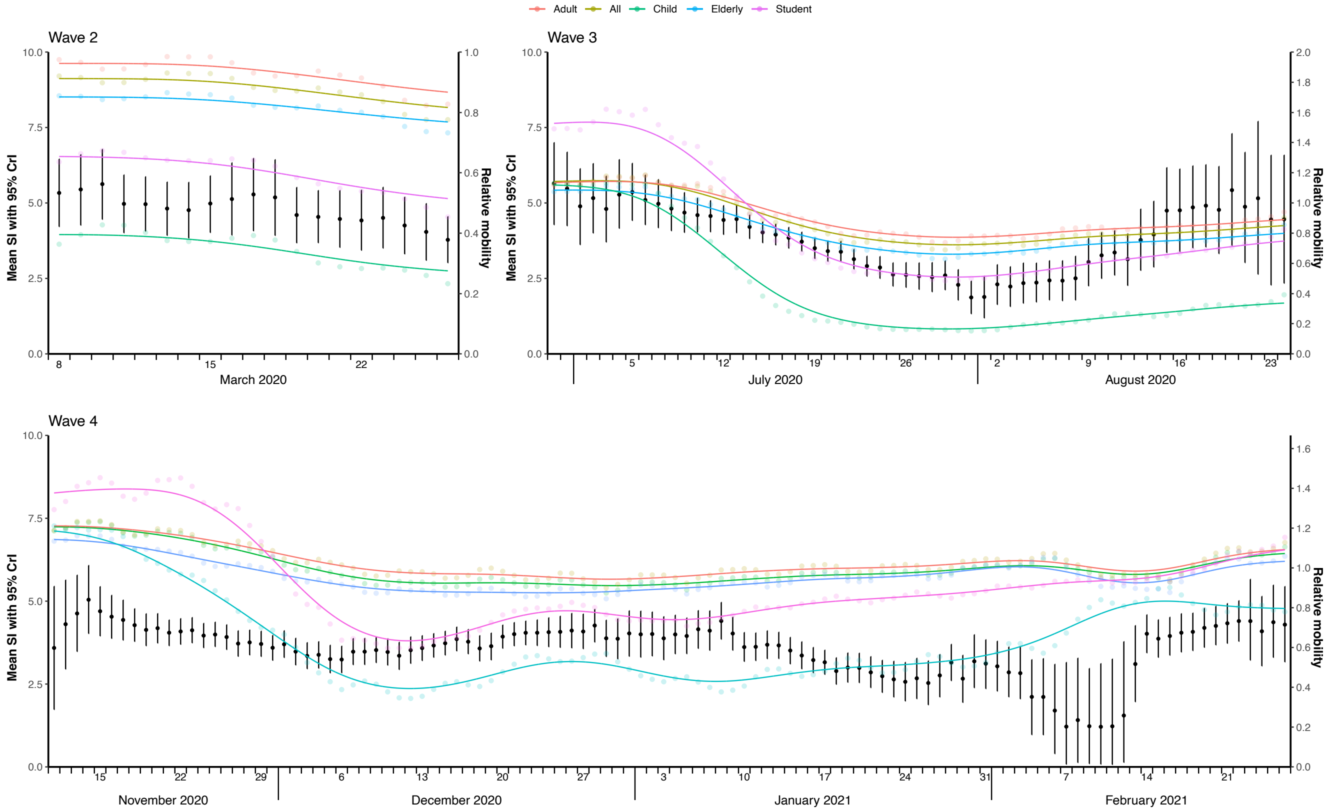

**Fig. S8. Effective serial interval estimates and relative mobility as the time series of digital travel-related Octopus card transactions during COVID-19 pandemic in Hong Kong.** The time varying estimates of effective serial intervals in black dots with 95% CrI (in black vertical line segments). The daily empirical relative mobility presented in the dots of respective colour for Children, Students, Adults, Elderly and All with their fitted lines (non-linear kernel regression) in respective colour, calculated based on the empirical data in 10-day sliding windows (presented with respect to mid-date as the 5th day of each sliding window).

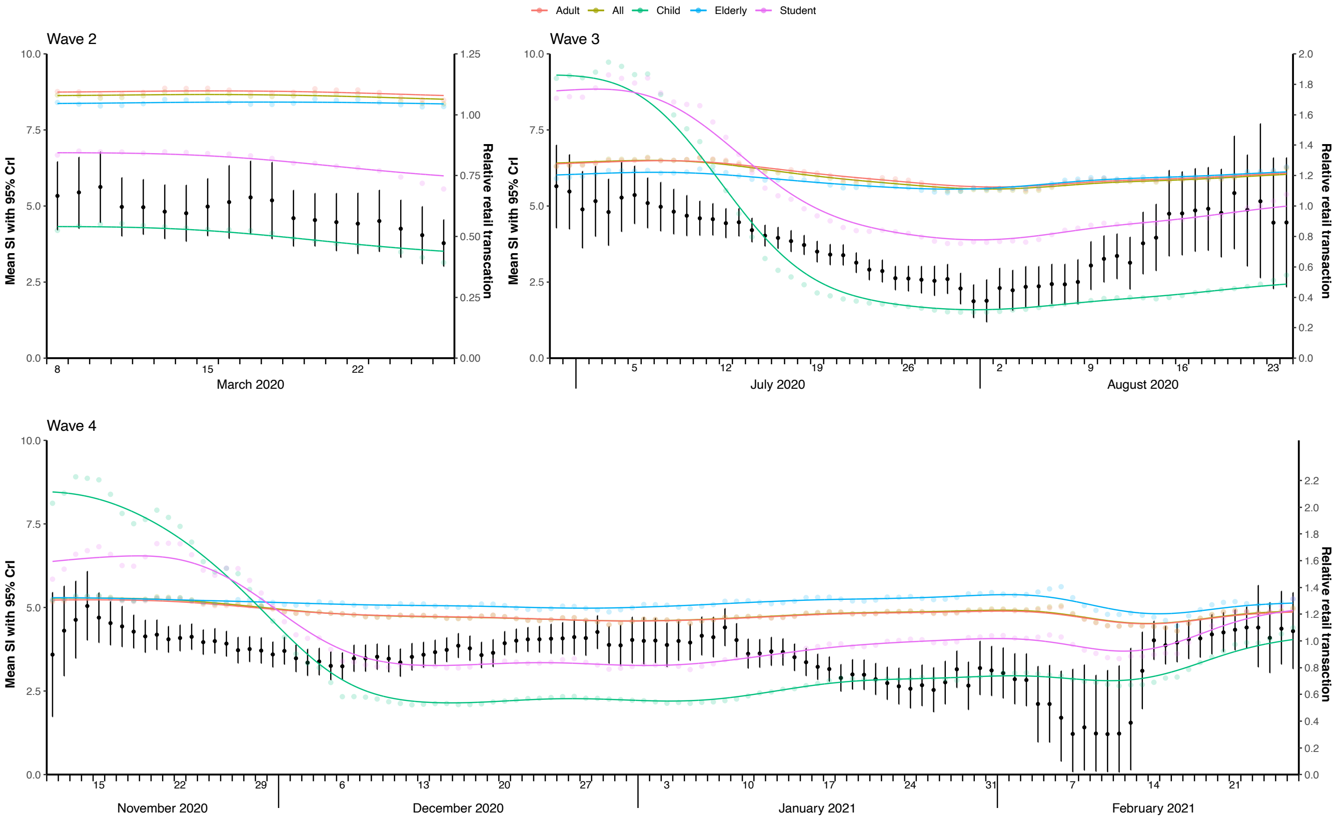

**Fig. S9. Effective serial interval estimates and relative retail transactions (mobility) as the time series of digital retail-related Octopus card transactions during COVID-19 pandemic in Hong Kong.** The time varying estimates of effective serial intervals in black dots with 95% CrI (in black vertical line segments). The daily empirical relative retail transactions **(**mobility) presented in the dots of respective colour for Children, Students, Adults, Elderly and All with their fitted lines (non-linear kernel regression) in respective colour, calculated based on the empirical data in 10-day sliding windows (presented with respect to mid-date as the 5th day of each sliding window).

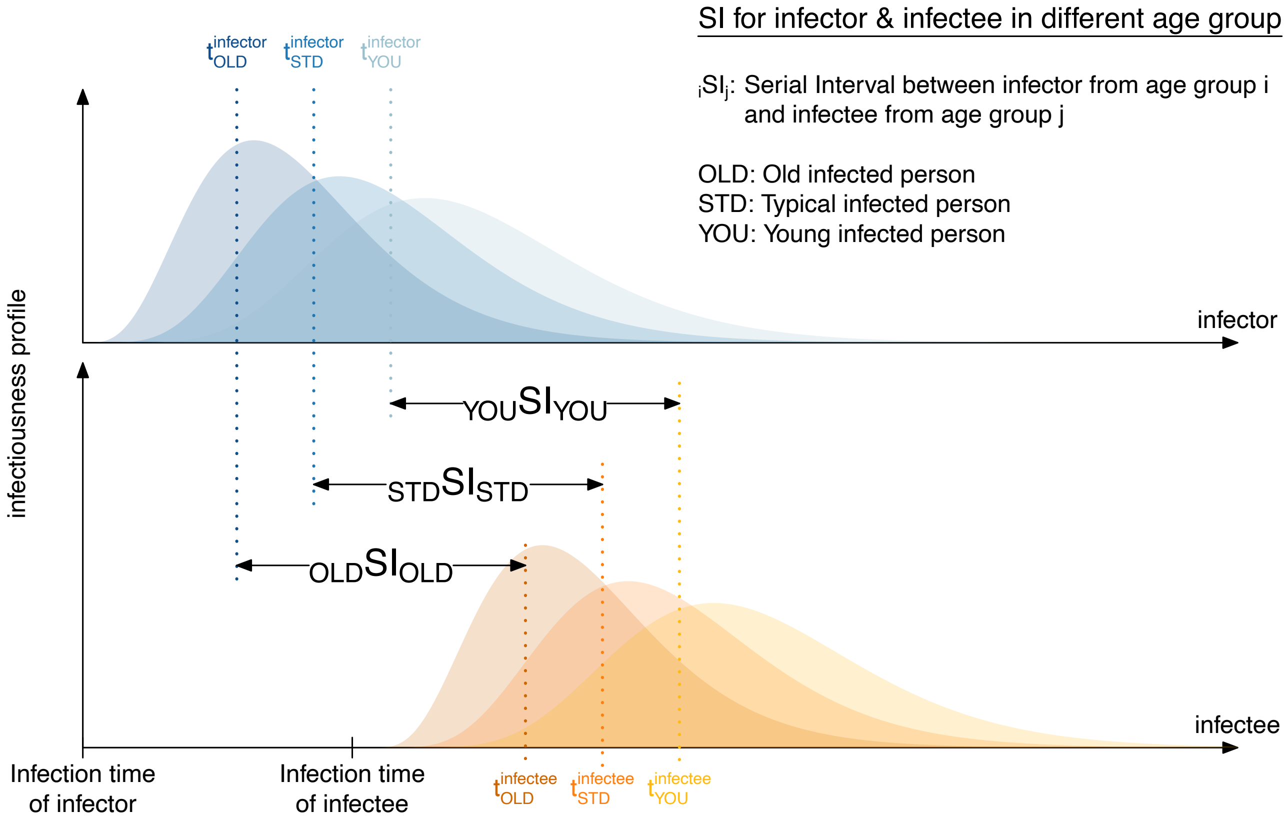

**Fig. S10. Illustration of the possible changes in the estimation of the serial interval distributions might be driven by the age of infector and infectee in the transmission chains.** The variation in viral load and severity may be related to the age of the infected individual, hence will have impact of latent period and incubation period, therefore may modulate the serial intervals accordingly. If the transmission chains consist of infector and infectee of same/similar age group, the mean serial intervals will be same (i.e., $_i SI_j$ for $i = j$), any other combination of the age/age-groups ($i \neq j$) of infetor and infectee will return the mean serial interval either shorter or longer based on $i > j$ or $i < j$ . The $i$ and $j$ are the age/age-group of infector and infectee. The same illustration can be extended for severity in general, accounting for other factors of severity and their respective impact on the viral load and infectiousness profile.

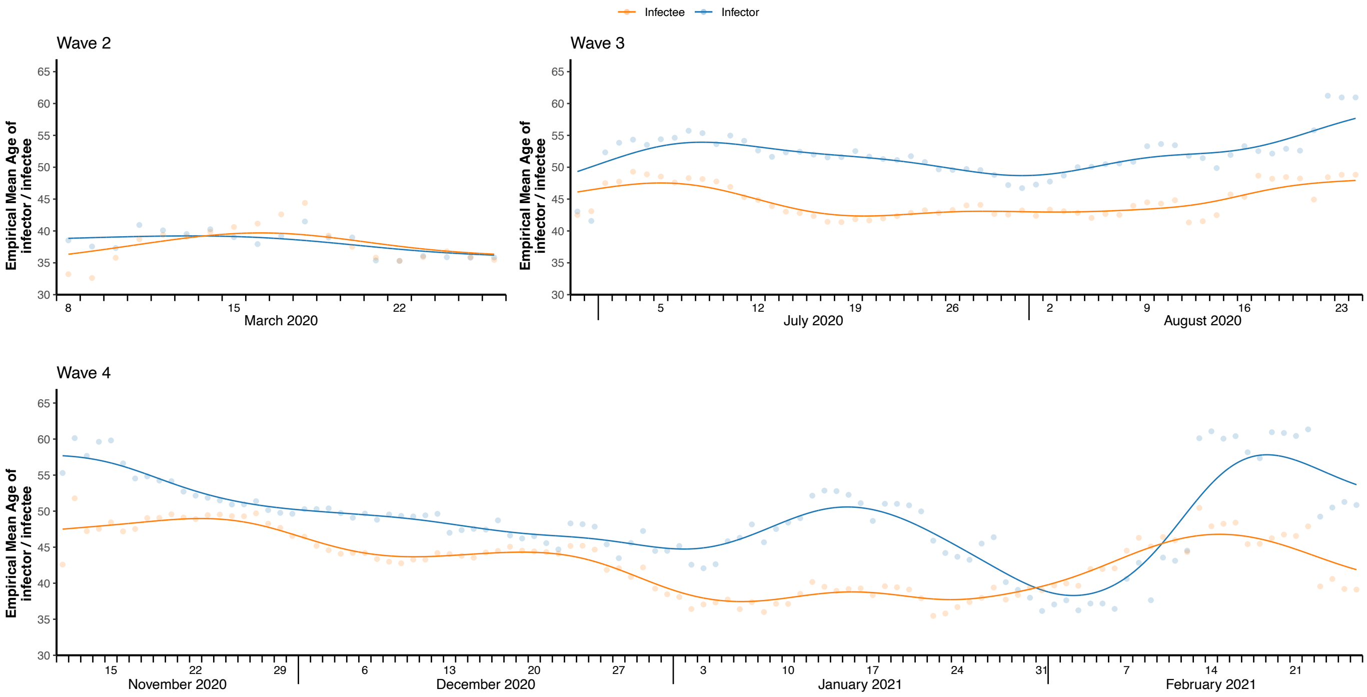

**Fig. S11. Temporal age-distributions of infectors and infectees in three waves of COVID-19 pandemic in Hong Kong.** The dots are empirical mean age for infectors (in light blue) and infectees (in light orange), with their fitted lines (non-linear kernel regression) in respective colour, calculated based on the empirical data in 10-day sliding windows (presented with respect to mid-date as the 5th day of each sliding window).

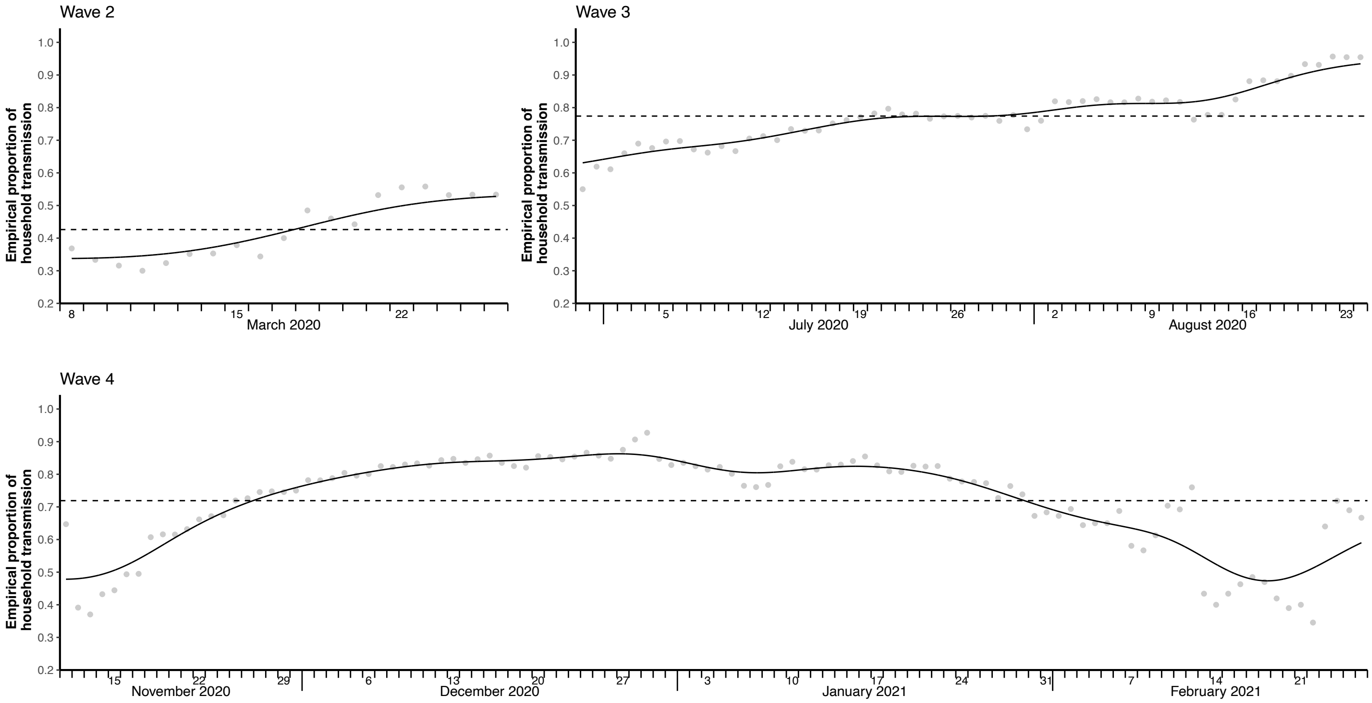

**Fig. S12. Temporal empirical proportion of household transmissions in three waves of COVID-19 pandemic in Hong Kong.** The grey dots are empirical proportion of household transmissions with their fitted lines (non-linear kernel regression) in black, calculated based on the empirical data in 10-day sliding windows (presented with respect to mid-date as the 5th day of each sliding window).

### 8. Supplementary Tables

**Table S1:** The summary of the public health and social measures (PHSMs) with the timing and duration implemented during four waves in Hong Kong.

| Waves | Dates | Types | subtype | tighten/ relax/ enhance | PHSMs | Links |
|---|---|---|---|---|---|---|
| 2 | 24/3/2020 | case | test | tighten | Test asymptomatic inbound travellers | https://www.news.gov.hk/eng/2020/03/20200324/20200324_231424_828.html?type=category&name=covid19&tl=t |
| 2 | 25/3/2020 | community | travel | tighten | Ban entry of non-Hong Kong residents from overseas countries/territories | https://www.news.gov.hk/eng/2020/03/20200323/20200323_164827_699.html?type=category&name=covid19&tl=t |
| **3** | 11/7/2020 | community | Social distancing | tighten | Bars and clubs gathering restriction from 8 to 4 | https://www.news.gov.hk/eng/2020/07/20200709/20200709_175812_722.html?type=category&name=covid19&tl=t |
| 3 | 15/7/2020 | community | social distancing | tighten | All leisure venues closure; group gathering no more than 4; restaurants stop dine-in/selling food from 6pm to 5 am | https://www.news.gov.hk/eng/2020/07/20200713/20200713_211216_022.html?type=category&name=covid19&tl=t; https://www.news.gov.hk/eng/2020/07/20200714/20200714_112235_478.html?type=category&name=covid19&tl=t; |
| 3 | 15/7/2020 | community | travel | tighten | Negative test result of travellers from high-risk areas required before entering HK | https://www.news.gov.hk/eng/2020/07/20200713/20200713_211216_022.html?type=category&name=covid19&tl=t |
| 3 | 17/7/2020 | case | test | tighten | Began testing high-risk occupation groups (start with taxi drivers and catering business) | https://www.news.gov.hk/eng/2020/07/20200717/20200717_223259_469.html?type=category&name=covid19&tl=t; https://www.news.gov.hk/eng/2020/07/20200716/20200716_202721_470.html?type=category&name=covid19&tl=t |
| 3 | 27/7/2020 | case | test | tighten | Start testing minibus drivers (free, voluntary) | https://www.news.gov.hk/eng/2020/07/20200726/20200726_131905_168.html?type=category&name=covid19 |
| 3 | 28/7/2020 | community | Social distancing | tighten | Chief Executive Carrie Lam urged public to stay at home, video released by government | https://www.news.gov.hk/eng/2020/07/20200728/20200728_231326_805.html?type=category&name=covid19&tl=t |
| 3 | 29/7/2020 | community | social distancing | tighten | Group gathering no more than 2 (relaxed to no more than 4 on Sep 11) | https://www.news.gov.hk/eng/2020/07/20200727/20200727_162631_406.html?type=category&name=covid19&tl=t |
| 3 | 11/8/2020 | case | test | tighten | Start testing ferry/MTR/tram operators (free, voluntary) | https://www.news.gov.hk/eng/2020/08/20200810/20200810_173103_863.html?type=category&name=covid19&tl=t |

| | | | | | | |
|---|---|---|---|---|---|---|
| 3 | 17/8/2020 | case | test | tighten | Start testing employees of bus companies and driving instructors (free, voluntary) | https://www.news.gov.hk/eng/2020/08/20200814/20200814_201936_205.html?type=category&name=covid19&tl=t |
| 3 | 20/8/2020 | case | test | tighten | Start testing more high-risk groups of people including supermarket staffs and postal office workers | https://www.news.gov.hk/eng/2020/08/20200820/20200820_192310_550.html?type=category&name=covid19&tl=t; https://www.news.gov.hk/chi/2020/08/20200820/20200820_101551_350.html?type=category&name=covid19&tl=t |
| 3 | 24/8/2020 | case | test | tighten | Start testing domestic helpers (free, voluntary) | https://www.news.gov.hk/eng/2020/08/20200823/20200823_153459_671.html?type=category&name=covid19&tl=t |
| 3 | 25/8/2020 | case | test | tighten | Start testing container terminal operators (free, voluntary) | https://www.news.gov.hk/eng/2020/08/20200825/20200825_224623_139.html?type=category&name=covid19&tl=t |
| 3 | 26/8/2020 | case | test | tighten | Start testing lift, escalator engineers (free, voluntary) | https://www.news.gov.hk/eng/2020/08/20200825/20200825_171209_456.html?type=category&name=covid19&tl=t |
| 3 | 28/8/2020 | case | test | tighten | Start testing frontline hotel staffs (free, voluntary) | https://www.news.gov.hk/eng/2020/08/20200827/20200827_230457_474.html?type=category&name=covid19&tl=t |
| 3 | 28/8/2020 | community | social distancing | relax | Extend dine-in in restaurants until 9 pm; reopen some of the leisure venues with little physical contacts | https://www.news.gov.hk/eng/2020/08/20200827/20200827_110426_985.html?type=category&name=covid19&tl=t |
| 3 | 1/9/2020 | community | Testing accessibility | enhance | Launch the Universal Community Testing Programme | https://www.news.gov.hk/eng/2020/08/20200828/20200828_131548_838.html?type=category&name=covid19&tl=t |
| 4 | 6/11/2020 | case | test | tighten | Test high-risk groups (bar staffs) from Nov 6 to Nov 13 (free, voluntary) | https://www.news.gov.hk/eng/2020/11/20201104/20201104_145519_442.html?type=category&name=covid19&tl=t |
| 4 | 9/11/2020 | case | test | tighten | Start testing school staffs | https://www.news.gov.hk/eng/2020/11/20201103/20201103_171356_118.html?type=category&name=covid19&tl=t |
| 4 | 13/11/2020 | case | quarantine | tighten | Mandatory 14-day hotel quarantine for overseas travellers | https://www.info.gov.hk/gia/general/202011/03/P2020110300613.htm |
| 4 | 15/11/2020 | case | test | tighten | Mandatory virus testing set | https://www.news.gov.hk/eng/2020/11/20201114/20201114_181652_667.html?type=category&name=covid19&tl=t |
| 4 | 16/11/2020 | community | Social distancing | tighten | Dine-in services suspended at midnight, maximum patrons from 8 to 4, Bar/pub restriction from 4 to 2 | https://www.news.gov.hk/eng/2020/11/20201114/20201114_181615_268.html?type=category&name=covid19&tl=t |
| 4 | 18/11/2020 | case | test | tighten | Start testing frozen food storage staffs | https://www.news.gov.hk/eng/2020/11/20201116/20201116_170338_194.html?type=category&name=covid19&tl=t |

| | | | | | | |
|---|---|---|---|---|---|---|
| 4 | 18/11/2020 | case | quarantine | tighten | Visiting people under compulsory hotel quarantine not allowed | https://www.news.gov.hk/eng/2020/11/20201114/20201114_181615_268.html?type=category&name=covid19&tl=t |
| 4 | 22/11/2020 | case | test | tighten | Mandatory testing for people visited specific dancing venues (HK had a huge dance club cluster COVID-19 cases) | https://www.news.gov.hk/eng/2020/11/20201122/20201122_172435_036.html?type=category&name=covid19&tl=t |
| 4 | 23/11/2020 | community | Testing accessibility | enhance | Mobile specimen collection stations provided | https://www.news.gov.hk/eng/2020/11/20201123/20201123_103518_203.html?type=category&name=covid19&tl=t |
| 4 | 24/11/2020 | case | test | tighten | Mandatory testing on people who had been to specific premises | https://www.news.gov.hk/eng/2020/11/20201124/20201124_105311_964.html?type=category&name=covid19&tl=t |
| 4 | 26/11/2020 | community | social distancing | tighten | Bars and clubs closed | https://www.news.gov.hk/eng/2020/11/20201124/20201124_174447_130.html?type=category&name=covid19&tl=t |
| 4 | 30/11/2020 | case | test | tighten | Mandatory test for care home staffs | https://www.news.gov.hk/eng/2020/12/20201201/20201201_102909_023.html |
| 4 | 2/12/2020 | community | social distancing | tighten | Group gathering no more than 2; school suspension | https://www.news.gov.hk/eng/2020/11/20201130/20201130_171503_290.html?type=category&name=covid19&tl=t; https://www.news.gov.hk/eng/2020/11/20201129/20201129_172437_678.html?type=category&name=covid19&tl=t |
| 4 | 8/12/2020 | case | test | tighten | Mandatory test 5 days after completion of quarantine | https://www.news.gov.hk/chi/2020/12/20201208/20201208_163125_181.html?type=category&name=covid19&tl=t |
| 4 | 9/12/2020 | case | test | tighten | Mandatory test on taxi drivers (Dec 9 - 22) | https://www.news.gov.hk/eng/2020/12/20201205/20201205_174005_553.html?type=category&name=covid19&tl=t |
| 4 | 10/12/2020 | community | Social distancing | tighten | Dine-in services suspended after 6pm | https://www.news.gov.hk/eng/2020/12/20201208/20201208_162805_968.html?type=category&name=covid19&tl=t |
| 4 | 12/12/2020 | case | test | tighten | Mandatory test on medical practitioners (Dec 12 - 25) | https://www.news.gov.hk/eng/2020/12/20201211/20201211_231212_258.html?type=category&name=covid19&tl=t |
| 4 | 24/12/2020 | case | quarantine | tighten | Extend quarantine period to 21-day hotel quarantine, and mandatory test on the 19th/20th days after arrival | https://www.news.gov.hk/eng/2020/12/20201224/20201224_231616_358.html?type=category&name=covid19&tl=t |
| 4 | 26/1/2021 | case | test | tighten | Restriction-testing strategy started (No one allowed to go out in targeted area until being tested negative) | https://www.news.gov.hk/eng/2021/01/20210126/20210126_201853_692.html?type=category&name=covid19&tl=t |
| 4 | 4/2/2021 | case | test | tighten | Mandatory test for HK international airport staffs (from Feb 4 to Feb 25) | https://www.news.gov.hk/eng/2021/02/20210202/20210202_141022_130.html?type=category&name=covid19&tl=t |
| 4 | 18/2/2021 | community | social distancing | relax | No more than 4 people can sit at one table in restaurants; Reopen recreational/leisure businesses and facilities; | https://www.news.gov.hk/eng/2021/02/20210216/20210216_182238_426.html?type=category&name=covid19&tl=t; https://www.news.gov.hk/eng/2021/02/20210210/20210210_175437_601.html?type=category&name=covid19&tl=t |

| | |
|---|---|
| | civil servants fully resume normal public services |

Notes: Travel-based PHSMs are often implemented as community-wide measures, hence included in the sub category of community-wide measures.

**Table S2:** Inter- and intra-wave (pre-, during and post-peak) estimates of serial interval and onset-to-isolation interval distributions (mean and standard deviation (sd) with 95% CrI) for COVID-19 in Hong Kong.

| Estimates | Periods | Wave 2 | | Wave 3 | | Wave 4 (1st Peak) | | Wave 4 (2nd Peak) | |
|---|---|---|---|---|---|---|---|---|---|
| | | Mean | sd | Mean | sd | Mean | sd | Mean | sd |
| **Serial Intervals** | Pre-peak | 5.5 (4.4, 6.6) | 2.4 (1.8, 3.5) | 4.6 (4.1, 5.0) | 3.9 (3.6, 4.2) | 4.0 (3.7, 4.4) | 2.8 (2.6, 3.1) | 4.0 (3.5, 4.5) | 3.0 (2.6, 3.4) |
| | During Peak | 4.5 (3.6, 5.4) | 3.2 (2.6, 4.0) | 3.1 (2.8, 3.5) | 3.2 (3.0, 3.5) | 3.5 (3.2, 3.8) | 3.1 (2.9, 3.3) | 3.1 (2.8, 3.5) | 2.7 (2.4, 3.0) |
| | Post-peak | 3.2 (1.9, 4.4) | 2.5 (1.8, 3.7) | 2.7 (2.2, 3.2) | 4.3 (3.9, 4.6) | 4.0 (3.6, 4.3) | 2.4 (2.2, 2.7) | 3.2 (2.7, 3.7) | 3.3 (3.0, 3.6) |
| **Onset-to-isolation intervals** | Pre-peak | 5.8 (4.8, 6.9) | 2.3 (1.8, 3.3) | 5.4 (5.0, 5.8) | 3.5 (3.2, 3.8) | 3.7 (3.4, 4.0) | 2.3 (2.1, 2.5) | 3.9 (3.5, 4.3) | 2.3 (2.1, 2.7) |
| | During Peak | 4.9 (3.6, 6.4) | 5.0 (4.0, 6.4) | 5.4 (5.1, 5.8) | 3.4 (3.1, 3.7) | 4.4 (4.1, 4.7) | 2.7 (2.6, 3.0) | 3.4 (3.1, 3.7) | 2.3 (2.1, 2.6) |
| | Post-peak | 3.5 (2.2, 5.2) | 3.0 (2.2, 4.5) | 5.2 (4.8, 5.6) | 3.5 (3.2, 3.8) | 4.6 (4.2, 4.9) | 2.6 (2.4, 2.9) | 5.0 (4.6, 5.5) | 2.8 (2.5, 3.2) |

**Table S3:** Regression models analysis of effective serial interval and public health and social measures (PHSMs) (including case isolation, other case-based and community-wide PHSMs).

| Periods | Models | Measures | | PHSMs at case | | | PHSMs at community | |
|---|---|---|---|---|---|---|---|---|
| | | | Intercept | Onset-to-isolation | Level-2 | Level-3 | Level-2 | Level-3 |
| Second wave | SI ~ Iso | Estimate (p-value) | 2.71 (<0.001) | 0.40 (<0.001) | | | | |
| | | $R^2$ $(df)$ | 0.60 (17) | | | | | |
| | SI ~ PHSMs* | Estimate (p-value) | 4.94 (<0.001) | | | | -0.91 (<0.001) | |
| | | $R^2$ $(df)$ | 0.49 (17) | | | | | |
| | SI ~ Iso + PHSMs* | Estimate (p-value) | 3.37 (<0.001) | 0.29 (0.004) | | | - 0.49 (0.040) | |
| | | $R^2$ $(df)$ | 0.70 (16) | | | | | |
| Third wave | SI ~ Iso | Estimate (p-value) | -0.58 (0.709) | 0.81 (0.006) | | | | |
| | | $R^2$ $(df)$ | 0.13 (55) | | | | | |
| | SI ~ PHSMs* | Estimate (p-value) | 4.90 (<0.001) | | | | -1.68 (<0.001) | -1.41 (<0.001) |
| | | $R^2$ $(df)$ | 0.39 (54) | | | | | |
| | SI ~ PHSMs* + Iso | Estimate (p-value) | 2.44 (0.10) | 0.44 (0.09) | | | - 1.56 (< 0.001) | -1.25 (<0.001) |
| | | $R^2$ $(df)$ | 0.42 (53) | | | | | |
| Fourth wave | SI ~ Iso | Estimate (p-value) | 3.96 (<0.001) | -0.10 (0.33) | | | | |
| | | $R^2$ $(df)$ | 0.01 (106) | | | | | |
| | SI ~ PHSMs Community | Estimate (p-value) | 4.16 (<0.001) | | | | -0.86 (<0.001) | |
| | | $R^2$ $(df)$ | 0.23 (106) | | | | | |
| | SI ~ PHSMs Case | Estimate (p-value) | 4.31 (<0.001) | | -0.68 (0.002) | -1.27 (<0.001) | | |
| | | $R^2$ $(df)$ | 0.24 (105) | | | | | |
| | SI ~ PHSMs Community + PHSMs Case | Estimate (p-value) | 4.31 (<0.001) | | 0.12 (0.625) | -0.61 (0.012) | -0.92 (<0.001) | |
| | | $R^2$ $(df)$ | 0.40 (104) | | | | | |
| | SI ~ PHSMs Community + PHSMs Case + Iso | Estimate (p-value) | 2.74 (<0.001) | 0.42 (< 0.001) | -0.08 (0.736) | -1.29 (< 0.001) | -0.86 (<0.001) | |
| | | $R^2$ $(df)$ | 0.49 (103) | | | | | |

Note: SI: serial interval (here effective serial interval), Iso: Onset-to-isolation interval; *: indicate the PHSMs for second and third waves were implemented almost simultaneously and unable disentangle for case-based and community-wide PHSMs as their proxies were with high temporal correlation. Level-i: indicates the respective level for the strength and timing of the PHSMs (case-based and community-wide) as implemented in Hong Kong (details in section 3 above). The case-based (targeted) PHSMs for fourth wave was mostly driven by different levels of testing strategy from individuals to mass, therefore indicating negative association in general.

**Table S4**: Interrupted univariate regression models analysis of effective serial interval and onset-to-isolation interval. The interrupted time-series were define based on the respective break points.

| **Periods and data-subsets** | | **Model results** | **Regression coefficients** | |
|---|---|---|---|---|
| | | | **Intercept** | **Onset-to-isolation** |
| Third wave | Subperiod-1 (Windows: Jul 07 – Jul 16, 2020 to Aug 07 – Aug 16, 2020) | Estimate (p-value) | 7.02 (0.009) | -0.75 (0.125) |
| | | $R^2$ $(df)$ | 0.08 (30) | |
| | Subperiod-2 (Other than subperiod-1) | Estimate (p-value) | 1.30 (0.075) | 0.63 (< 0.001) |
| | | $R^2$ $(df)$ | 0.53 (23) | |
| Fourth wave | Subperiod-1 (Windows: Nov 07 – Nov 16, 2020 to Nov 22 – Dec 01, 2020) | Estimate (p-value) | 2.32 (0.182) | 0.52 (0.261) |
| | | $R^2$ $(df)$ | 0.09 (14) | |
| | Subperiod-2 (Windows: Nov 23 – Dec 02, 2020 to Dec 18 – Dec 27, 2020) | Estimate (p-value) | 2.81 (0.002) | 0.18 (0.327) |
| | | $R^2$ $(df)$ | 0.04 (24) | |
| | Subperiod-3 (Windows: Dec 19 – Dec 28, 2020 to Jan 17 – Jan 26, 2021) | Estimate (p-value) | 0.26 (0.638) | 0.93 (< 0.001) |
| | | $R^2$ $(df)$ | 0.60 (28) | |
| | Subperiod-4 (Windows: Jan18 – Jan 27, 2021 to Feb 13 – Feb 22, 2021) | Estimate (p-value) | 0.83 (0.362) | 0.36 (0.057) |
| | | $R^2$ $(df)$ | 0.14 (25) | |
| | Subperiod-5 (Windows: Feb 14 – Feb 23, 2021 to Feb 22 – Mar 03, 2021) | Estimate (p-value) | 4.35 (< 0.001) | -0.02 (0.795) |
| | | $R^2$ $(df)$ | 0.01 (7) | |

**Table S5:** Regression models analysis of effective serial interval and public health and social measures (PHSMs). The relative population mobility (based on Octopus card transactions) and per capita PCR testing volumes in Hong Kong during the epidemic waves (for 2nd, 3rd and 4th) of the COVID-19 pandemic assumed to reflect the measures of PHSMs.

| **Periods** | **Models** | **Octopus Mobility (or uses)** | | | | | | **Per Capita Testing Volume (Per 10,000 population)** | | |
|---|---|---|---|---|---|---|---|---|---|---|
| | | **Travel Related** | | | **Retail Related** | | | | | |
| | | Intercept (p-value) | Coeff (p-value) | $R^2$ $(df)$ | Intercept (p-value) | Coeff (p-value) | $R^2$ $(df)$ | Intercept (p-value) | Coeff (p-value) | $R^2$ $(df)$ |
| Second wave | SI ~ Child | 2.23 (<0.001) | 7.44 (<0.001) | 0.81 (17) | 0.32 (0.565) | 8.94 (<0.001) | 0.79 (17) | | | |
| | SI ~ Students | 0.78 (0.078) | 6.73 (<0.001) | 0.85 (17) | -2.62 (0.014) | 9.18 (<0.001) | 0.78 (17) | | | |
| | SI ~ Adults | -1.88 (0.207) | 7.20 (<0.001) | 0.56 (17) | -10.38 (0.211) | 13.90 (0.075) | 0.18 (17) | | | |
| | SI ~ Elderly | -2.94 (0.051) | 9.43 (<0.001) | 0.64 (17) | 4.01 (0.735) | 0.75 (0.947) | 0.00 (17) | | | |
| | SI ~ All | -1.97 (0.150) | 7.71 (<0.001) | 0.61 (17) | -11.12 (0.175) | 14.79 (0.059) | 0.19 (17) | 5.96 (<0.001) | -0.42 (<0.001) | 0.82 (17) |
| Third wave | SI ~ Child | 2.80 (<0.001) | 2.33 (<0.001) | 0.54 (52) | 2.84 (<0.001) | 1.34 (<0.001) | 0.48 (52) | | | |
| | SI ~ Students | 2.01 (<0.001) | 2.17 (<0.001) | 0.57 (52) | 1.39 (<0.001) | 2.21 (<0.001) | 0.56 (52) | | | |
| | SI ~ Adults | -1.70 (0.009) | 6.07 (<0.001) | 0.59 (52) | -12.79 (<0.001) | 13.77 (<0.001) | 0.63 (52) | | | |
| | SI ~ Elderly | -0.79 (0118) | 5.66 (<0.001) | 0.62 (52) | -20.32 (<0.001) | 20.55 (<0.001) | 0.69 (52) | | | |
| | SI ~ All | -0.87 (<0.109) | 5.36 (<0.001) | 0.59 (52) | -11.65 (<0.001) | 12.92 (<0.001) | 0.66 (52) | 6.95 (<0.001) | -0.24 (<0.001) | 0.73 (52) |
| Fourth wave | SI ~ Child | 3.02 (<0.001) | 0.81 (0.014) | 0.06 (106) | 2.99 (<0.001) | 0.62 (<0.001) | 0.14 (106) | | | |
| | SI ~ Students | 2.68 (<0.001) | 0.92 (0.005) | 0.07 (106) | 2.52 (<0.001) | 0.96 (<0.001) | 0.11 (106) | | | |
| | SI ~ Adults | 0.74 (0.438) | 2.72 (0.004) | 0.08 (106) | -0.57 (0.733) | 3.42 (0.015) | 0.05 (106) | | | |
| | SI ~ Elderly | 1.98 (0.035) | 1.63 (0.093) | 0.03 (106) | 6.33 (0.003) | -2.17 (0.189) | 0.02 (106) | | | |
| | SI ~ All | 1.19 (0.159) | 2.35 (0.006) | 0.07 (106) | -0.20 (0.899) | 3.10 (0.018) | 0.05 (106) | 6.21 (<0.001) | -0.11 (<0.001) | 0.31 (92) |

Note: SI: serial interval (here effective serial interval). For 4th wave, we excluded the data beyond 12 February, 2022 as during this period most of the transmission pairs are mostly from K11 restaurant cluster, which was limited under control from community transmission.

**Table S6:** Factor-specific estimates of serial interval distributions with the mean and standard deviation (sd) based on the confirmed and likely pairs with the onset intervals between the infector and the infectees within 8 days for the three epidemic waves in Hong Kong. Numbers in the brackets show the estimates when the pairs with the onset interval between 5 days and 14 days were included in the analysis. 'NA' indicates no or insufficient transmission pairs data to estimate serial interval distribution.

| **Factors** | | **Second wave** | | **Third wave** | | **Fourth wave** | |
|---|---|---|---|---|---|---|---|
| | | Mean | sd | Mean | sd | Mean | sd |
| **Age** | Below 65 | 4.5 (4.4, 4.9) | 2.9 (2.9, 3.2) | 3.3 (2.7, 3.8) | 3.8 (3.8, 4.2) | 3.5 (2.8, 4.1) | 2.9 (2.7, 3.5) |
| | Above 65 | NA | NA | 4.1 (3.4, 4.6) | 3.9 (3.9, 4.3) | 4.1 (3.4, 5.5) | 2.9 (2.8, 4.0) |
| **Sex** | Male | 4.6 (4.4, 5.2) | 3.0 (3.0, 3.3) | 3.3 (2.7, 3.8) | 3.8 (3.7, 4.1) | 3.6 (2.9, 4.5) | 2.7 (2.4, 3.5) |
| | Female | 4.3 (4.3, 4.4) | 2.9 (2.9, 3.0) | 3.6 (2.9, 4.1) | 3.9 (3.9, 4.3) | 3.6 (2.9, 4.4) | 3.1 (3.0, 3.8) |
| **Setting** | Household | 4.8 (4.6, 5.0) | 3.1 (3.1, 3.2) | 3.6 (2.9, 4.1) | 3.8 (3.8, 4.2) | 3.6 (2.9, 4.3) | 2.9 (2.7, 3.5) |
| | Non-household | 4.2 (4.1, 4.9) | 2.8 (2.8, 3.3) | 3.1 (2.5, 3.6) | 3.9 (3.8, 4.2) | 3.8 (3.2, 5.0) | 3.0 (2.9, 4.0) |
| **Outcome** | Critical | 5.4 (5.4, 5.4) | 4.9 (4.9, 4.9) | 4.5 (3.9, 5.1) | 4.1 (4.1, 4.3) | 4.0 (3.1, 6.0) | 2.9 (2.8, 4.4) |
| | Serious | NA | NA | 3.8 (3.2, 4.1) | 3.3 (3.2, 3.5) | 3.8 (3.1, 4.5) | 2.8 (2.5, 3.5) |
| | Stable | 4.2 (4.1, 4.8) | 2.6 (2.6, 3.0) | 3.3 (2.7, 3.8) | 3.8 (3.8, 4.2) | 3.6 (3.0, 4.3) | 2.8 (2.6, 3.5) |
| **Onset-to-isolation delay** | Shorter | 3.7 (3.7, 3.9) | 2.5 (2.4, 2.6) | 2.6 (2.0, 2.9) | 3.4 (3.3, 3.8) | 3.2 (2.4, 3.8) | 3.0 (2.9, 3.6) |
| | Longer | 5.5 (5.5, 6.2) | 3.2 (3.2, 3.5) | 4.6 (4.0, 5.2) | 4.1 (4.1, 4.4) | 4.5 (3.5, 5.6) | 2.6 (2.4, 3.5) |

**Table S7**: Identifying the significant factors of serial interval estimation by Kruskal Wallis test (p-values) based on the confirmed pairs plus the additional likely pairs under pre-defined threshold of 8 day.

| **Factors** | **Comparison groups** | **2nd wave** | | **3rd wave** | | **4th wave** | | **Pooled** | |
|---|---|---|---|---|---|---|---|---|---|
| | | p-value | Sample size | p-value | Sample size | p-value | Sample size | p-value | Sample size |
| **Age** | Below 65 yrs, Above 65 yrs | NA | 87, 0 | 0.023 | 759, 206 | 0.001 | 1088, 293 | < 0.001 | 1934, 499 |
| **Sex** | Male, Female | 0.400 | 53, 34 | 0.120 | 478, 487 | 0.834 | 710, 671 | 0.505 | 1241, 1192 |
| **Setting** | Household, Non-household | 0.353 | 42, 45 | 0.037 | 746, 219 | 0.614 | 1072, 309 | 0.236 | 1860, 573 |
| **Outcome** | Critical, Serious | 0.115 | 8, 2 | 0.537 | 84, 82 | 0.1831 | 95, 80 | 0.235 | 187, 164 |
| | Critical, Stable | 0.600 | 8, 77 | 0.037 | 84, 796 | 0.022 | 95, 1190 | 0.002 | 187, 2063 |
| | Serious, Stable | 0.022 | 2, 77 | 0.201 | 82, 796 | 0.680 | 80, 1190 | 0.202 | 164, 2063 |
| **Onset-to-isolation delay** | Shorter, Longer | 0.005 | 52, 35 | < 0.001 | 555, 410 | < 0.001 | 902, 479 | < 0.001 | 1390, 1043 |

**Table S8:** Biases in using single constant serial interval throughout the wave over effective serial interval distributions in three waves of COVID-19 in Hong Kong. The biases are measured by attack rates (epidemic sizes) and peak intensity and comparing them as evaluated under different counterfactual serial interval (SI) distributions (pre-peak, peak, and post-peak timing) for second, third and fourth waves (with two peaks).

| Incidences and peak intensities as observed and predicted using counterfactual SI* | 2nd wave | | | 3rd wave | | | 4th wave | | |
|---|---|---|---|---|---|---|---|---|---|
| | Total number of cases | Peak cases (daily) | Bias as reduction / increase (%) | Total number of cases | Peak cases (daily) | Bias as reduction / increase (%) | Total number of cases | Peak cases (daily) | Bias as reduction / increase (%) |
| **Observed** | **212** | **20** | | **2738** | **131** | | **4016** | **93** | |
| Effective SI | 272 | 20 | + 28% | 2911 | 131 | + 6% | 4056 | 93 | + 0.1% |
| Pre-peak SI | 393 | 39 | + 85% | 2344 | 117 | - 14% | 1225 | 29 | - 69% |
| During peak SI | 51 | 4 | - 76% | 433 | 16 | - 84% | 244 | 5 | - 94% |
| Post-peak SI | 9 | 1 | - 96% | 143 | 5 | - 95% | 1645 | 39 | - 59% |
| Pre-peak SI (2nd peak) | | | | | | | 974 | 23 | - 76% |
| During peak SI (2nd peak) | | | | | | | 174 | 3 | - 96% |
| Post-peak SI (2nd peak) | | | | | | | 114 | 2 | - 97% |

Notes: 2nd wave: onset cases from March 4 to April 8, 2020, 3rd wave: onset cases from June 25 to September 8, 2020, 4th wave: onset cases from November 1, 2020 to March 23, 2021, SI: Serial interval. * Counterfactual predicted incidences and peak intensities are based on the reconstruction of epi-curves using effective SI and counterfactual (pre-, during and post-peak) single constant serial intervals.
